# Contextual factors influencing implementation of tuberculosis digital adherence technologies: a scoping review guided by the RE-AIM framework

**DOI:** 10.1101/2024.06.16.24308969

**Authors:** Shruti Bahukudumbi, Chimweta I. Chilala, Nicola Foster, Barbie Patel, Mona S. Mohamed, Miranda Zary, Cedric Kafie, Genevieve Gore, Kevin Schwartzman, Katherine Fielding, Ramnath Subbaraman

**Affiliations:** Department of Public Health and Community Medicine and Center for Global Public Health, Tufts University School of Medicine, Boston, United States; TB Centre, London School of Hygiene and Tropical Medicine, London, United Kingdom; McGill International Tuberculosis Centre, Research Institute of the McGill University Health Centre, Montréal, Canada; Schulich Library of Physical Sciences, Life Sciences, and Engineering; McGill University, Montréal, Canada; Division of Geographic Medicine and Infectious Diseases, Tufts Medical Center, Boston, United States

## Abstract

**Introduction:** Digital adherence technologies (DATs) may enable person-centered tuberculosis (TB) treatment monitoring; however, implementation challenges may undermine their effectiveness. Using the RE-AIM framework, we conducted a scoping review to identify contextual factors informing “reach” (DAT engagement by people with TB) and “adoption” (DAT uptake by healthcare providers or clinics).

**Methods:** We searched eight databases from January 1, 2000 to April 25, 2023 to identify all TB DAT studies. After extracting qualitative and quantitative findings, using thematic synthesis, we analyzed common findings to create meta-themes informing DAT reach or adoption. Meta-themes were further organized using the Unified Theory of Acceptance and Use of Technology, which posits technology use is influenced by perceived usefulness, ease of use, social influences, and facilitating conditions.

**Results:** 66 reports met inclusion criteria, with 61 reporting on DAT reach among people with TB and 27 reporting on DAT adoption by healthcare providers. Meta-themes promoting reach included perceptions that DATs improved medication adherence, facilitated communication with providers, made people feel more “cared for,” and enhanced convenience compared to alternative care models (perceived usefulness); and lowered stigma (social influences). Meta-themes limiting reach included literacy and language barriers and DAT technical complexity (ease of use); increased stigma (social influences); and suboptimal DAT function and complex cellular accessibility challenges (facilitating conditions). Meta-themes promoting adoption included perceptions DATs improved care quality or efficiency (perceived usefulness). Meta-themes limiting adoption included negative DAT impacts on workload or employment and suboptimal accuracy of adherence data (perceived usefulness); and suboptimal DAT function, complex cellular accessibility challenges, and insufficient provider training (facilitating conditions). Limitations of this review include the limited studies informing adoption meta-themes.

**Conclusion:** This review identifies diverse contextual factors that can inform improvements in DAT design and implementation to achieve higher engagement by people with TB and healthcare providers, which could improve intervention effectiveness.

**KEY MESSAGES:** *What is already known on this topic:*

- Digital adherence technologies (DATs) are increasingly used to monitor TB treatment; however, systematic reviews suggest DATs have mixed effectiveness for improving TB outcomes and suboptimal accuracy for measuring medication adherence.
- Inadequate DAT “reach” (engagement by people with TB) and “adoption” (uptake by healthcare providers) may contribute to their limited effectiveness and accuracy.
- Understanding contextual factors influencing DAT reach and adoption may be critical to improve the design, implementation, and public health impact of TB DATs.

*What this study adds:*

- Our findings show people with TB value DATs when they improve adherence, enhance communication with providers, enhance convenience of care, and reduce stigma.
- People with TB are less likely to engage with DATs in settings with barriers to cellular accessibility or when DATs are not designed for their literacy level, are technically complex, have suboptimal function, or increase stigma.
- TB healthcare providers value DATs when they improve care quality or efficiency.
- Healthcare providers are less likely to engage in settings with barriers to cellular accessibility or when DATs increase workloads, threaten employment, provide inaccurate adherence data, or have suboptimal function.

*How this study might affect research, practice, or policy:*

- Our findings may inform future design of DATs to focus on what people with TB value, such as improved communication with providers and convenience of care.
- Our findings may also help to identify settings in which DATs are unlikely to be effective, such as locations where cellular accessibility barriers are substantial due to poor infrastructure.

## INTRODUCTION

Digital adherence technologies (DATs) have been increasingly studied and implemented as part of routine TB care over the last decade.[1,2] These technologies—which include interventions involving short messaging service (SMS) texting, feature (non-smart) phones, video-supported therapy (VST), digital pillboxes, and ingestion sensors—provide reminders and enable electronic observation of dose ingestion, often with the aim of replacing in-person direct observation of therapy (DOT).

Despite this enthusiasm, evaluations of the evidence regarding TB DATs reveal challenges limiting their public health impact.[3,4] For example, a recent systematic review evaluating the effectiveness of DATs for improving TB treatment outcomes revealed mixed findings.[5] Although some DATs, like VST, were associated with improved treatment outcomes in high- and upper-middle-income countries, most technologies evaluated in low- or lower-middle-income countries, which account for most people with TB globally, were not associated with improved outcomes. Another recent systematic review evaluated the performance of DATs for measuring TB medication adherence, because an important function of some DATs is identification of nonadherent individuals who may need additional support.[6] This review found DATs often have variable or suboptimal performance for measuring adherence in real-world conditions.

The limited effectiveness and accuracy of TB DATs may be explained, in part, by implementation challenges. RE-AIM (Reach, Effectiveness, Adoption, Implementation, and Maintenance) is a framework for assessing implementation outcomes and contextual factors.[7] Two RE-AIM outcomes are of particular relevance to this paper. “Reach” refers to the proportion of people who receive an intended public health intervention; we use reach to refer to the proportion of people taking TB treatment who receive or engage with DAT interventions. “Adoption” refers to the proportion of healthcare providers or settings that engage in delivering a public health intervention; we use adoption to refer to the proportion of healthcare providers or clinics appropriately delivering or engaging with DAT interventions.

A forthcoming scoping review evaluating DAT implementation outcomes shows that serial drop-offs in DAT engagement by people with TB—e.g., suboptimal cellphone access, provision of DATs to people with TB, or initial or sustained engagement by people with TB—contribute to poor overall reach of DATs in many settings, especially low- and lower-middle-income countries.[8] Although findings on adoption were more limited, the review also found high variability in DAT adoption across clinics in some settings. While that review quantified implementation outcomes, its findings did not attempt to provide insights into contextual barriers contributing to suboptimal reach and adoption.

In this paper, which is a companion to the DAT implementation outcomes scoping review, we report findings of a scoping review aimed at identifying contextual factors informing reach and adoption of DATs. We conducted a scoping review, rather than a systematic review, given the heterogeneity of study types reporting contextual findings and the flexibility that a scoping review allows in combining different findings using thematic synthesis.[9] We further organized findings using constructs in the Unified Theory of Acceptance and Use of Technology (UTAUT), which helps to understand and predict technology engagement.[10] By understanding contextual barriers, this review may inform the design and implementation of future DAT interventions so as to enhance their reach, adoption, effectiveness, and public health impact.

## METHODS

### PICOS framework and scoping review design

The review was conceived as a systematic review and registered in PROSPERO, the International Prospective Register of Systematic Reviews (CRD42022326968). During data extraction, based on the heterogeneity in study designs, we modified this to a scoping review, though we followed the original protocol except that we did not assess study quality or risk of bias. We followed the Preferred Reporting Items for Systematic Reviews and Meta-Analyses Extension for Scoping Reviews (PRISMA-ScR) guideline (checklist is in online supplementary appendix 1).[11]

Our study design is guided by a Population, Intervention, Comparison, Outcomes, and Study design (PICOS) framework.[12] For the RE-AIM reach outcome, the population includes people being treated for TB disease or infection, including children with TB, people with drug-resistant TB, and people with HIV. For the RE-AIM adoption outcome, the population includes healthcare providers, program managers, or policymakers involved in DAT implementation. The intervention comprises TB DATs, defined as an intervention with a digital component (which could be part of a multi-component intervention) intending to measure or promote adherence, reduce missed clinic visits, or reduce loss to follow-up. Studies may or may not have included a comparison group. When reported, comparison groups usually comprised various DOT approaches (e.g., clinic-based DOT, family DOT) or self-administered therapy. Outcomes included all RE-AIM dimensions; however, we only report findings on DAT reach and adoption, given limited contextual findings on other RE-AIM dimensions.[7] We extracted qualitative contextual findings (e.g., emergent themes from interviews or focus groups) and quantitative contextual findings from structured surveys (e.g., percentage of people reporting specific benefits of DATs).

### Search strategy

Initial and refresher searches without language restrictions were conducted by a librarian, collectively spanning January 1, 2000 to April 25, 2023 (updated from April 14, 2022), in MEDLINE/Ovid, Embase, the Cochrane Central Register of Controlled Trials, CINAHL, Web of Science, clinicaltrials.gov, and Europe PMC to capture preprints (including those in medRxiv). As any TB DAT study could report contextual findings on implementation, our search strategy was broad, including only two concepts with related terms: “tuberculosis” (including TB disease and infection) and “digital adherence technologies,” including terms for specific TB DATs (the full search strategy is in online supplementary appendix 1). Finally, we extracted and screened all references from systematic reviews identified in our initial search to assess the validity of our search and identify outstanding studies. We found no further studies meeting the inclusion criteria.

### Inclusion and exclusion criteria

We included studies if they reported on DAT implementation (not hypothetical use) during treatment for TB disease or infection and if they reported contextual findings on at least one RE-AIM outcome, excluding effectiveness. Although data collection may have been embedded within other study designs (e.g., cohort studies or randomized trials), relevant contextual findings usually involved qualitative interviews, structured surveys, or direct observation of people with TB or healthcare providers. After extracting data, sufficient contextual findings were only available for RE-AIM outcomes of reach and adoption, though the RE-AIM implementation outcome (i.e., the fidelity with which providers adhere to an intervention protocol) is indirectly informed by our review findings as described later.

We excluded protocols, review articles, editorials, and commentaries; articles in which the technology was not used to measure or improve adherence; and articles in which clinical or implementation outcomes were reported without contextual findings.

### Screening strategy and study selection

After de-duplication using EndNote (Version 20.2.1, Clarivate, London, United Kingdom), two reviewers (among SB, CC, MS, MZ, CK and NF) independently screened titles and abstracts in a blinded manner using Rayyan.ai (Cambridge, United States). Full text articles were independently assessed in a blinded manner for inclusion by two reviewers, with conflicts resolved by senior investigators (RS, KF, or KS).

### Data extraction

Data from included studies were extracted into an Excel template by five reviewers (SB, CC, NF, KF, and RS). Given the large number of articles, for each article, one reviewer extracted data and a second verified these data, with conflicts resolved through discussion or involvement of a third reviewer. We extracted data on study characteristics, design, setting (inpatient or outpatient), participant characteristics, type of DAT, and details on the intervention program in which the DAT was embedded. We extracted contextual findings into separate Excel sheets for RE-AIM outcomes of reach, adoption, implementation, and maintenance, though findings on the latter two outcomes were extremely limited. We separated qualitative and quantitative contextual findings and included separate columns for each UTAUT construct (described further below).

### Data analysis: synthesis of meta-themes and reporting using the UTAUT

After completing data extraction, authors SB and RS engaged in an iterative process using thematic synthesis to create new “meta-themes” that may inform increased or suboptimal reach or adoption. Thematic synthesis is an extension of meta-ethnography originally described by Noblit and Hare for conducting qualitative systematic reviews,[9,13,14] an approach that has previously been used for TB qualitative systematic reviews.[15] Our approach to thematic synthesis involved aggregating similar themes into a new meta-theme that reflects a broader concept from the underlying findings. More specifically, author SB copied extracted findings onto a new Excel spreadsheet, which organized findings by whether they informed increased or suboptimal reach. Within increased or suboptimal reach, new sheets were created to organize themes by each UTAUT construct. SB and RS met on a weekly basis over a few months to iteratively group similar themes together and then create meta-themes to reflect broader underlying concepts, with routine feedback from CC, NF, and KF. The same process was followed for adoption.

Our thematic synthesis approach is unique in that we included both qualitative and quantitative data as contextual findings, as there was similarity in concepts across both types of data. For a quantitative finding to be included in the synthesis, it had to be reported by at least 15% of people surveyed to indicate that it had meaningful influence on reach or adoption. For adverse effects of DATs or concerns related to stigma, privacy, or confidentiality only, we included quantitative findings if reported by at least 5% of people surveyed, as such challenges could be associated with significant harm (e.g., disclosure of TB diagnosis). Given our scoping review approach, we did not evaluate or exclude studies based on quality, and this is actually consistent with the approach taken by many qualitative systematic reviews.[15]

Final meta-themes were reported using the UTAUT, which describes the following constructs that influence technology use: performance expectancy, effort expectancy, social influences, and facilitating conditions.[10] Performance expectancy, or perceived usefulness, is the extent to which a person thinks the DAT helps with TB care (for people with TB) or care delivery (for healthcare providers). Effort expectancy, or ease of use, refers to how easily people are able to navigate and use the DAT. Social influences refers to the role of other individuals (e.g., family members) or societal factors in influencing DAT use. Facilitating conditions refers to the quality of institutional support or infrastructure to ensure success of the DAT intervention. Throughout this paper, we use “perceived usefulness” and “ease of use” to refer to performance expectancy and effort expectancy, respectively, given the more intuitive meaning of the former terms.

## RESULTS

### Characteristics of the included studies

Of 14,416 unique abstracts identified through the search, 771 underwent full-text review, of which 66 had relevant data for extraction (figure 1). Of the 66 included articles, 61 (92%) reported contextual findings on reach of DATs among people with TB, while 27 (41%) reported on adoption of DATs by healthcare providers. Of the 66 included articles, the primary DAT was SMS in 14 studies, feature (non-smart) phone-based approaches (including 99DOTS) in 10 studies, VST in 24 studies, digital pillboxes in 12 studies, ingestion sensors in 2 studies, and App-based in 5 studies. 99DOTS is a feature phone-based DAT that requires people with TB to call a number every day to report medication ingestion.[16]

**Figure 1.**
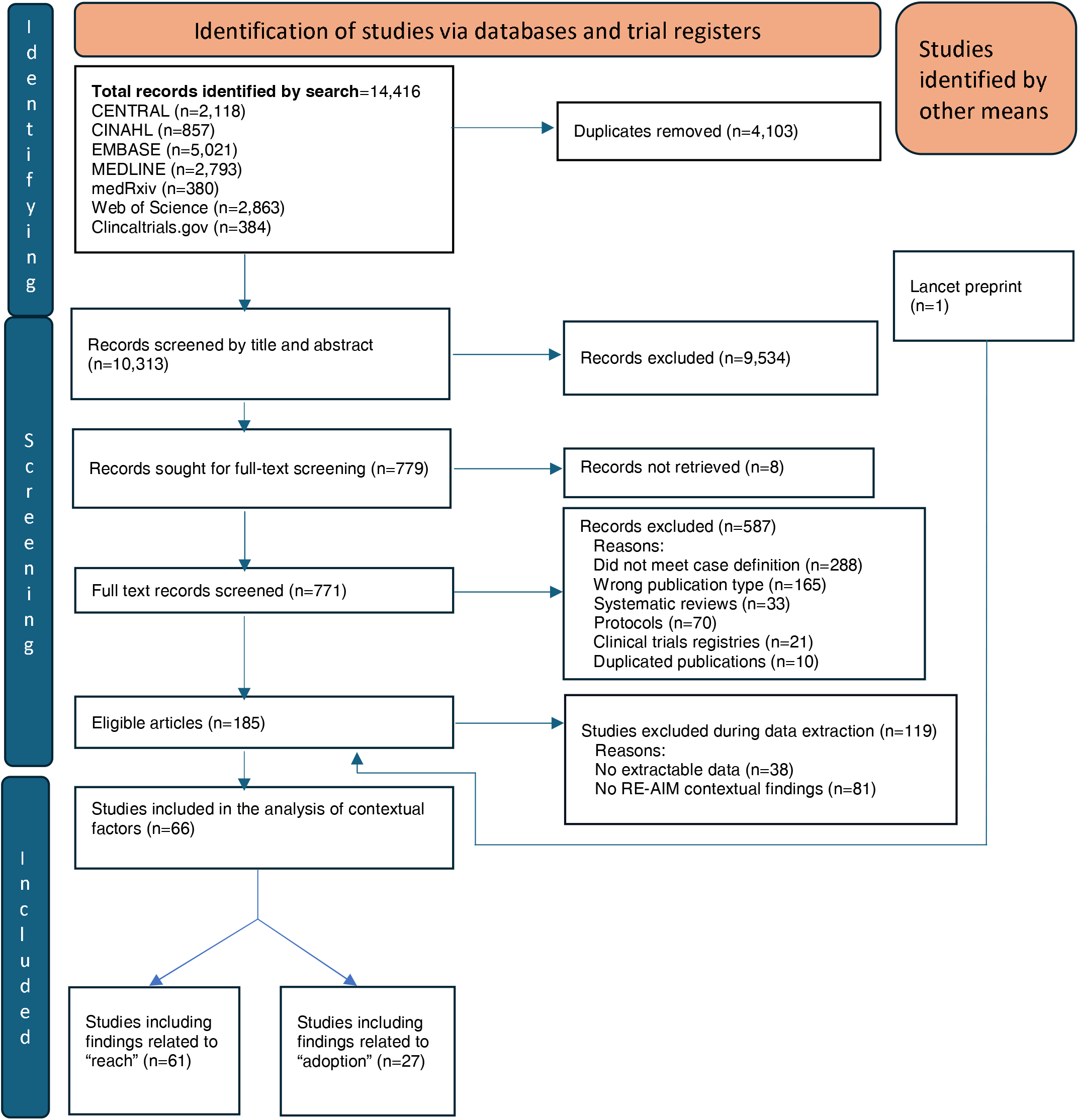
PRISMA flow diagram summarizing the process of identification of included studies

### Contextual factors promoting reach of DATs among people with TB

With regard to the UTAUT construct of perceived usefulness, four meta-themes related to the perceptions that DATs enhanced the ability of people with TB to engage in treatment: improved medication adherence behavior, better storage and organization of medications (specifically in relation to digital pillboxes), enhanced access to information and knowledge, and better monitoring of medication adverse effects (Table 2; findings informing each meta-theme for reach are provided in online supplementary appendix 2). With regard to improved medication adherence behavior, which was the most frequently reported of these themes, people with TB reported that reminders at consistent times “trained” [their] minds” to develop a new “awareness of pill-taking.”[75]

**Table 1.** Characteristics of studies included in this scoping review evaluating contextual factors implementation of TB digital adherence technologies.

| Author Year | Dominant DAT modality | Type of TB | Countries | Country income level | RE-AIM dimension (i.e., reach, adoption, or both) | Type of data reported (i.e., qualitative, quantitative or both) |
| --- | --- | --- | --- | --- | --- | --- |
| Bardosh 2017 [17] | Phone | TB Infection | Canada, Kenya | High, Lower middle | Both | Both |
| Bassett 2016 [18] | SMS | Drug-sensitive TB | South Africa | Upper middle | Reach | Quantitative |
| Bediang 2018 [19] | SMS | Drug-sensitive TB | Cameroon | Lower middle | Reach | Quantitative |
| Belknap 2013 [20] | Ingestion Sensor | Drug-sensitive TB | United States | High | Reach | Quantitative |
| Bendiksen 2020 [21] | VST | Drug-sensitive TB | Norway | High | Both | Both |
| Bionghi 2018 [22] | Digital Pillbox | Drug-resistant TB | South Africa | Upper middle | Reach | Both |
| Bommakanti 2020 [23] | VST | Drug-sensitive TB | United States | High | Reach | Quantitative |
| Browne 2019 [24] | Ingestion Sensor | Drug-sensitive TB | United States | High | Reach | Quantitative |
| Buchman 2017 [25] | VST | Drug-sensitive TB | United States | High | Reach | Qualitative |
| Burzynski 2022 [26] | VST | Drug-sensitive TB | United States | High | Reach | Quantitative |
| Chen 2020 [27] | VST | TB Infection | Taiwan, China | High | Reach | Quantitative |
| Chuck 2016 [28] | VST | Drug-sensitive TB | United States | High | Reach | Quantitative |
| Cox 2018 [29] | SMS | Drug-sensitive TB | India | Lower middle | Reach | Quantitative |
| Cross 2019 [16] | 99DOTS | Drug-sensitive TB | India | Lower middle | Reach | Qualitative |
| Daftary 2017 [30] | Phone | TB Infection | Ethiopia | Low | Both | Both |
| Das Gupta 2020 [31] | Phone | Drug-sensitive TB | India | Lower middle | Reach | Qualitative |
| de Sumari-de Boer 2016 [32] | Digital Pillbox | Drug-sensitive TB | Tanzania | Lower middle | Reach | Qualitative |
| DeMaio 2001 [33] | VST | Drug-sensitive TB | United States | High | Reach | Quantitative |
| Dessie Gashu 2020 [34] | Phone | Drug-sensitive TB | Ethiopia | Low | Reach | Both |
| Drabarek 2019 [35] | Digital Pillbox | Drug-sensitive TB | Vietnam | Lower middle | Both | Qualitative |
| Garfein 2020 [36] | VST | Drug-sensitive TB | United States | High | Reach | Quantitative |
| Garfein 2015 [37] | VST | Drug-sensitive TB | United States, Mexico | High, Upper middle | Reach | Quantitative |
| Gashu 2021 [38] | SMS | Drug-sensitive TB | Ethiopia | Low | Reach | Qualitative |
| Gassanov 2013 [39] | VST | Drug-sensitive TB | Canada | High | Reach | Qualitative |
| Getachew 2022 [40] | Digital Pillbox | Drug-sensitive TB | Ethiopia | Low | Adoption | Both |
| Guo 2020 [41] | VST | Drug-sensitive TB | China | Upper middle | Reach | Quantitative |
| Guo 2020 [42] | VST | Drug-sensitive TB | China | Upper middle | Reach | Quantitative |
| Hermans 2017 [43] | SMS | Drug-sensitive TB | Uganda | Low | Reach | Both |
| Hirsch-Moverman 2017 [44] | SMS | Drug-sensitive TB | South Africa | Upper middle | Both | Both |
| Hoffman 2010 [45] | VST | Drug-sensitive TB | Kenya | Lower middle | Both | Both |
| Holzman 2018 [46] | VST | Drug-sensitive TB | United States | High | Both | Both |
| Holzman 2019 [47] | VST | Drug-sensitive TB | India | Lower middle | Reach | Quantitative |
| Horter 2014 [48] | App-based | Drug-resistant TB | United Kingdom, Australia, Philippines, Swaziland, Central African Republic, Uganda, South Africa, India, Armenia | High, High, Lower middle | Both | Qualitative |
| Iribarren 2020 [49] | App-based | Drug-sensitive TB | Argentina | Upper middle | Reach | Qualitative |
| Iribarren 2015 [50] | SMS | Drug-sensitive TB | Argentina | Upper middle | Both | Both |
| Iribarren 2013 [51] | SMS | Drug-sensitive TB | Argentina | Upper middle | Both | Both |
| Kalita 2021 [52] | Phone | Drug-sensitive TB | India | Lower middle | Reach | Both |
| Khachadourian 2020 [53] | SMS | Drug-sensitive TB | Armenia | Upper middle | Reach | Quantitative |
| Kopanitsa 2017 [54] | App-based | Drug-sensitive TB | Russian Federation | Upper middle | Both | Qualitative |
| Krueger 2010 [55] | VST | Drug-sensitive TB | United States | High | Adoption | Quantitative |
| Lam 2018 [56] | VST | TB Infection | United States | High | Both | Quantitative |
| Liu 2015 [57] | SMS, Digital Pillbox | Drug-sensitive TB | China | Upper middle | Reach | Quantitative |
| Mahmud 2010 [58] | SMS | Drug-sensitive TB | Malawi | Low | Both | Both |
| Milligan 2021 [59] | App-based | Drug-sensitive TB | Argentina | Upper middle | Both | Qualitative |
| Mohammed 2016 [60] | SMS | Drug-sensitive TB | Pakistan | Lower middle | Reach | Quantitative |
| Mohammed 2012 [61] | SMS | Drug-sensitive TB | Pakistan | Lower middle | Reach | Both |
| Moulding 2002 [62] | Digital Pillbox | Drug-sensitive TB | Haiti | Lower middle | Adoption | Quantitative |
| Navin 2017 [63] | App-based | Drug-sensitive TB | India | Lower middle | Reach | Quantitative |
| Nhavoto 2017 [64] | SMS | Drug-sensitive TB | Mozambique | Low | Reach | Both |
| Olano-Soler 2017 [65] | VST | Drug-sensitive TB | United States | High | Both | Both |
| Person 2011 [66] | Phone | TB Infection | United States | High | Reach | Qualitative |
| Prabhu 2021 [67] | 99DOTS | Drug-sensitive TB | India | Lower middle | Reach | Qualitative |
| Ratchakit-Nedsuwan 2020 [68] | Digital Pillbox | Drug-sensitive TB | Thailand | Upper middle | Reach | Qualitative |
| Ritter 2017 [69] | VST | Drug-sensitive TB | United States | High | Adoption | Qualitative |
| Sinkou 2017 [70] | VST | Drug-sensitive TB, Drug-resistant TB | Belarus | Upper middle | Reach | Qualitative |
| Stagg 2020 [71] | Digital Pillbox | Drug-sensitive TB | China | Upper middle | Reach | Qualitative |
| Story 2019 [72] | VST | Drug-sensitive TB | United Kingdom | High | Both | Quantitative |
| Thekkur 2019 [73] | 99DOTS | Drug-sensitive TB | India | Lower middle | Both | Qualitative |
| Thomas 2021 [74] | Digital Pillbox | Drug-resistant TB | India | Lower middle | Both | Qualitative |
| Thomas 2020 [75] | 99DOTS | Drug-sensitive TB | India | Lower middle | Both | Qualitative |
| Ting 2020 [76] | VST | Drug-sensitive TB | Australia | High | Reach | Qualitative |
| Trajman 2010 [77] | Digital Pillbox | TB Infection | Canada, Brazil, Saudi Arabia | High, Upper middle, High | Both | Qualitative |
| van den Boogaard 2011 [78] | Digital Pillbox | Drug-sensitive TB | Tanzania | Lower middle | Reach | Quantitative |
| Wade 2012 [79] | VST | Drug-sensitive TB | Australia | High | Both | Qualitative |
| Wade 2009 [80] | VST | Drug-sensitive TB | Australia | High | Reach | Qualitative |
| Wang 2019 [81] | Digital Pillbox | Drug-sensitive TB | China | Upper middle | Adoption | Quantitative |

App, application; DAT, digital adherence technology; RE-AIM, Reach, Effectiveness, Adoption, Implementation, Maintenance; SMS, short messaging service; TB, tuberculosis; VST, video-supported therapy.

**Table 2.** Meta-themes promoting increased reach (i.e., DAT use by people with TB) with findings organized using the UTAUT.

| Emerging meta-themes divided by UTAUT construct | Number of studies reporting meta-theme [citations] | DATs for which finding was reported (Number of studies per DAT) | Representative quotation on the meta-theme |
| --- | --- | --- | --- |
| **Perceived usefulness** |  |  |  |
| Improved medication adherence behavior | 23 [16,19,22,27,30–32,34,37,41,42,44–46,51,53,54,64,68,70,74,75,78] | SMS (6), Phone-based (4), 99DOTS (2), Digital pillbox (4), VST (6), App-based (1) | “[E]very time I forgot to take my medication, I was getting a reminder SMS then I find myself developing the habit of taking my pills...” (SMS)<br>“I have a new awareness that I should take tablets at 11 o’ clock. If we forget to call, the message alerts us...So it trains our mind to take pills on time...” (99DOTS) [1] |
| Better storage and organization of medications | 3 [22,32,74] | Digital pillbox (3) | “There are different compartments for each tablet, so they don’t get mixed...I like the arrows with the dots that explain how many of each medication I need to take.” (Digital pillbox) [74] |
| Enhanced access to information and knowledge | 5 [45,49,51,59,64] | SMS (2), VST (1), App-based (2) | “...if you didn’t know where to look for information, this way you have it at hand [in application] and you do not have to go look for it in books...” (App-based) [49] |
| Better monitoring of medication adverse effects | 5 [41,49,52,59,65] | Phone-based (1), VST (2), App-based (2) | Description of treatment supporter follow-up by App of adverse effects reported by a person with TB: “I saw that you reported nausea and vomiting. Is it a stomachache? Tell me when you are taking your medication” (App-based) [59] |
| Facilitated communication with providers | 12 [17,30,44,49–51,58,59,63–65,68] | SMS (5), Phone-based (2), Digital pillbox (1), VST (1), App-based (3) | “[I]t helps me to call and communicate with them when I am sick and receive calls from the clinic. Rather than totally depend on somebody, I can receive calls and I am able to call them when appropriate.” (Phone-based) [30] |
| Improved relationship between person with TB and provider | 4 [59,74,75,79] | 99DOTS (1), Digital pillbox (1), VST (1), App-based (1) |  |
| Fostered a feeling of being “cared for” | 11 [16,17,22,30,38,44,51,61,74,75,78] | SMS (4), Phone-based (2), 99DOTS (2), Digital pillbox (3) | “If I don’t open the [pill]box, somebody from the health center calls me to find out whether I have taken the tablets or not. They care for me.” (Digital pillbox) [74]<br>“It made me feel very important to have people worried about how I take my medication.” (Digital pillbox) [22] |
| Convenience of care delivery | 5 [17,43,46,50,79] | SMS (2), Phone-based (1), VST (2) | “The flexibility [of the technology] is good. You can be texting a patient. They have a problem. You go and find that [specialist] in the clinic. You call and give the phone right to the health worker. They get immediate feedback!” (Phone-based) [17] |
| Convenience compared to alternative care models | 20 [17,21,24,27,30,33, | SMS (1), Phone-based (2), 99DOTS (1), Digital pillbox (3), VST (13) | “Coming every day to the hospital is definitely more difficult than learning to connect to a conference on whatever a |
|  | 37,39,41,45,46,51,68,70,72,74–76,79,80] |  | laptop, tablet. I mean, it's pretty flexible..." (VST) [76] |
| Improved quality of care | 5 [17,45,46,50,75] | SMS (1), Phone-based (1), 99DOTS (1), VST (2) |  |
| **Ease of use** |  |  |  |
| DAT was easy to use | 13 [20,22,23,33,36,37,41,42,49,54,63,64,76] | SMS (1), Digital pillbox (1), Ingestion sensors (1), VST (8), App-based (3) |  |
| DAT provided flexibility and choice (e.g., timing of observation, monitoring approach) | 3 [26,65,79] | VST (3) |  |
| **Social influences** |  |  |  |
| Empowerment or enhanced autonomy | 5 [46,48,51,53,54] | SMS (2), VST (1), App-based (2) | "Through blogging you can share thoughts without thinking of what others will think. It helps to write what happened and what my experience was..." (App-based) [48] |
| Lower or reduced stigma from DAT use | 14 [21,22,27,33,36,37,39,41,43,44,46,68,74,79] | SMS (2), Digital Pillbox (3), VST (9) | "[P]eople undermine and discriminate you when you are on [HIV antiretroviral therapy], so the device is very helpful because nobody can see what is inside." (Digital pillbox) [22] |
| Positive family involvement in care or social support | 7 [18,36,48,52,53,74,75] | SMS (1), Phone-based (2), 99DOTS (1), Digital pillbox (1), VST (1), App-based (1) | "[My son] taught me, 'You have to take 2 tablets per day and follow the arrow mark from the starting point'...He gave me my medicines from the beginning [of therapy] and reminds me to take tablets...he also dials the toll-free numbers for me." (99DOTS) [75] |
| **Facilitating conditions** |  |  |  |
| Adequate DAT training by providers enabled engagement | 2 [37,74] | Digital pillbox (1), VST (1) |  |
| Provision of cellphone enabled DAT engagement | 1 [36] | VST (1) |  |
| Optimal DAT function or DAT problems easily resolved | 4 [37,57,65,79] | SMS (1), VST (3) |  |
| No or only minor challenges with network function | 2 [37,79] | VST (2) |  |

App, Application; DAT, digital adherence technology; SMS, short messaging service; TB, tuberculosis; UTAUT, Unified Theory of Acceptance and Use of Technology; VST, video-supported therapy. 99DOTS is a phone-based DAT.

Three meta-themes within perceived usefulness pertained to the impact of DATs on relationships between people with TB and their healthcare providers: facilitated communication with providers, improved relationship between the person with TB and providers, and fostered a feeling of being cared for. Of these, facilitated communication with providers was most frequently reported; people with TB valued having timely access to providers to ask questions about adverse effects, TB transmission risk, and appointment scheduling.[17,30,44,49–51,58,59,63–65,68] Several studies reported that DATs fostered a sense among people with TB of being “cared for” or “seen” by the health system because of an additional point of connection through technology.[16,17,22,30,38,44,51,61,68,74,75]

Three meta-themes within perceived usefulness pertained to perceptions of convenience and quality of care: convenience of care delivery, convenience compared to alternative care models, and improved quality of care. People with TB most frequently reported improved convenience of care in relation to more restrictive care models, such as facility-based DOT, in which people must visit clinics multiple times a week for observation of dosing. In contrast, DATs minimized life disruptions by reducing clinic visits, providing flexibility in dosing timings, and increasing treatment accessibility for people in rural areas or with demanding work schedules.[17,21,24,27,30,33,37,39,41,42,45,46,51,68,70,72,74–76,79,80]

With regard to the UTAUT construct of ease of use, two meta-themes emerged: the DAT was easy to use and the DAT provided flexibility and choice. People appreciated DATs for which there were few difficulties in tasks such as sending SMS texts or recording videos. People also valued flexibility in how and when they could report doses taken, for example when VST interventions allowed for asynchronous options.[26,65,79]

With regard to the UTAUT construct of social influences, three meta-themes emerged: empowerment or enhanced autonomy, low or reduced stigma from DAT use, and positive family involvement in care or social support. In several studies, people with TB experienced lower stigma due to perceptions of higher privacy and confidentiality of digital observation of dosing, usually in comparison to more restrictive in-person DOT models.[21,22,27,33,36,37,39,41,43,44,46,68,74,79]

With regard to the UTAUT construct of facilitating conditions, four meta-themes emerged, suggesting that DAT engagement by people with TB could be supported by adequate training in the DAT by healthcare providers, provision of cellphones (for phone-based DATs), provision DATs that function optimally or for which problems are easily resolvable, and settings with high-functioning cellular networks.

### Contextual factors limiting reach of DATs among people with TB

Several meta-themes emerged related to the UTAUT construct of perceived usefulness, of which five had findings from three or more studies: DAT does not address patient problems, technology fatigue, different or modified DAT modality desired, negative impact on relationship with healthcare system, and preference for in-person healthcare provider-patient communication (Table 3; findings informing each meta-theme for reach are provided in online supplementary appendix 2). Some people with TB conveyed dissatisfaction with DATs, because the technologies inadequately addressed critical problems contributing to nonadherence, such as medication adverse effects, alcohol use, or drug stockouts.[17,31,34] People with TB also described technology fatigue, as they felt overwhelmed by frequent notifications and texting or calling requirements, resulting in “decreased interest”[75] and motivation to engage with the DAT.[51,67,73,75]

**Table 3.**
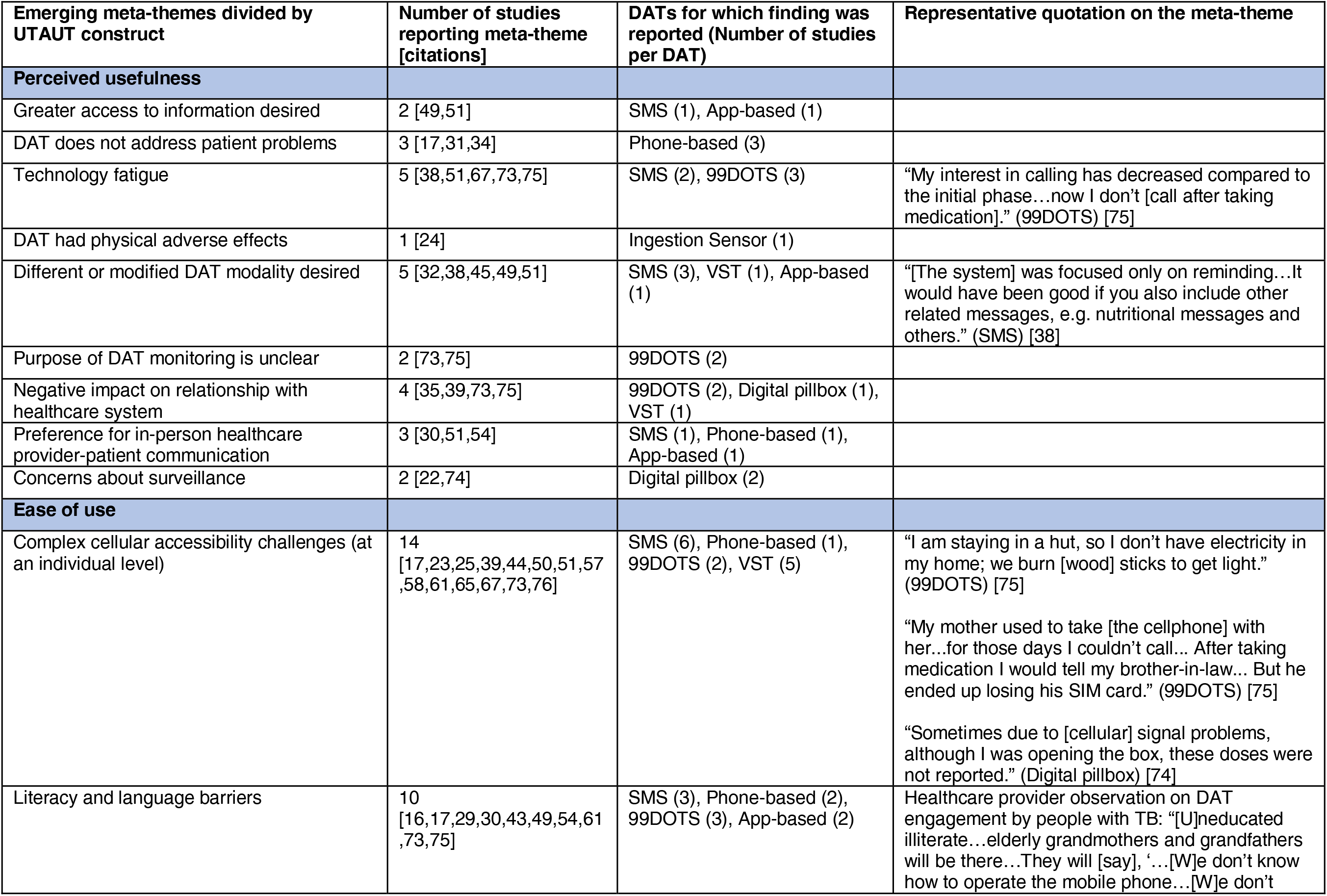
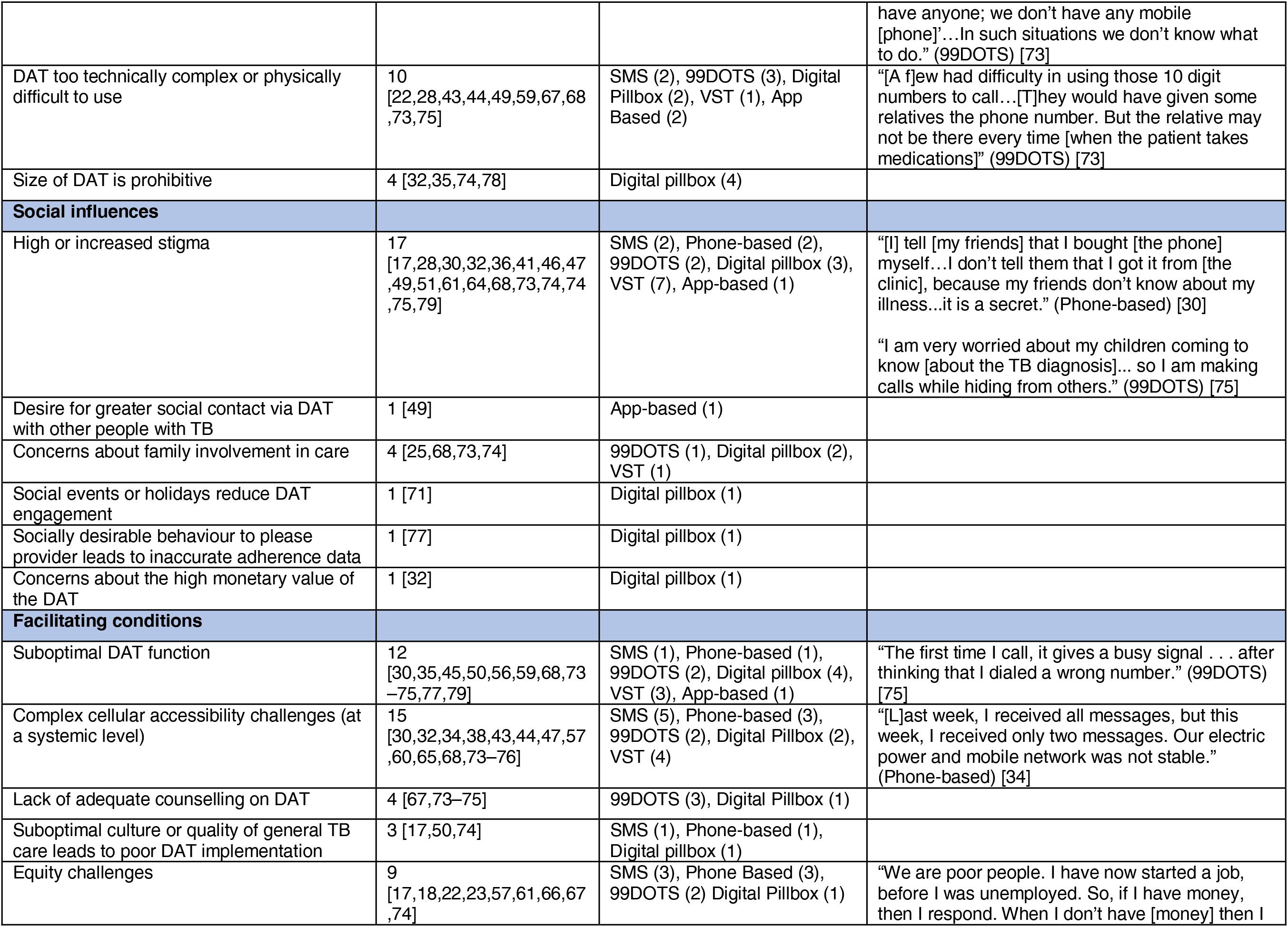
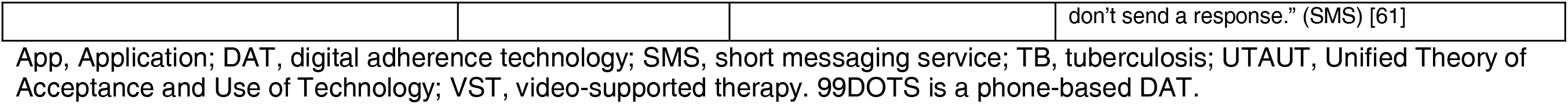
Meta-themes limiting reach (i.e., DAT use by people with TB) with findings organized using the UTAUT.

With regard to the UTAUT construct of ease of use, meta-themes had findings from several studies, suggesting that these barriers strongly limit reach of some DATs. People with TB reported numerous challenges to cellular accessibility at an individual level, including lack of access to cellphones, shared cellphone use within the family, running out of credit, challenges refilling credit, and lack of a stable cellular signal or internet connection.[17,23,25,39,44,50,51,57,58,61,65,67,73,76] Literacy and language barriers were also diverse and included SMS texts sent in foreign languages, struggles comprehending technical language, and reliance on family members to interpret messages.[16,17,29,30,43,49,54,61,73,75]

With regard to the UTAUT construct of social influences, the most frequently reported meta-themes were high or increased stigma and concerns about family involvement in care. Stigma-related concerns often resulted from the increased visibility of pill-taking with DATs. For example, people with TB reported privacy concerns from having to make a phone or video call to report dose ingestion and that receiving a new phone or digital pillbox (especially with audible reminders) marked them as having TB or HIV.[17,28,30,32,36,41,46,47,49,51,61,64,68,73–75,79]

With regard to the UTAUT construct of facilitating conditions, the most frequently reported meta-themes were suboptimal DAT function, complex cellular accessibility challenges at a systemic level, and equity challenges. Suboptimal DAT function included problems such as cellphone calls not connecting (for 99DOTS);[73,75] loud reminder alarms, malfunctioning reminder lights, or discrepant reminder timings (for digital pillboxes);[35,68,74,77] device battery failures; and difficulties accessing a DAT App.[30,45,50,56,59,79] Cellular accessibility challenges at a systemic level included inconsistent power supply and weak or inconsistent wireless fidelity (Wi-Fi) or cellular signals.[30,32,34,38,43,44,47,57,60,65,68,73–76] Equity challenges included differences in the ability of people with TB to access or engage with DATs by sex, age, educational status, income level, and urban versus rural location.[17,18,22,23,57,61,66,67,75]

### Contextual factors promoting DAT adoption by healthcare providers

With regard to the UTAUT construct of perceived usefulness, meta-themes with the most findings promoting adoption were: improved quality of care for people with TB, improved communication between providers and people with TB, better efficiency of care delivery, and improved data quality (Table 4; findings informing each meta-theme for adoption are provided in online supplementary appendix 3). Healthcare providers valued DATs when they improved quality of care, through improved adherence monitoring, response to questions from people with TB, or detection of medication adverse effects (reported as a separate meta-theme).[44–46,58,65,75,79,81] Improved efficiency of care delivery due to DATs manifested as time savings and reduced transportation costs from not conducting in-person DOT and an ability to care of more people per provider, which led to reductions in staff, alleviation of clinic crowding, or increased clinic capacity.[21,40,44,46,54,55,58,65,69,72,74,75,79]

**Table 4.**
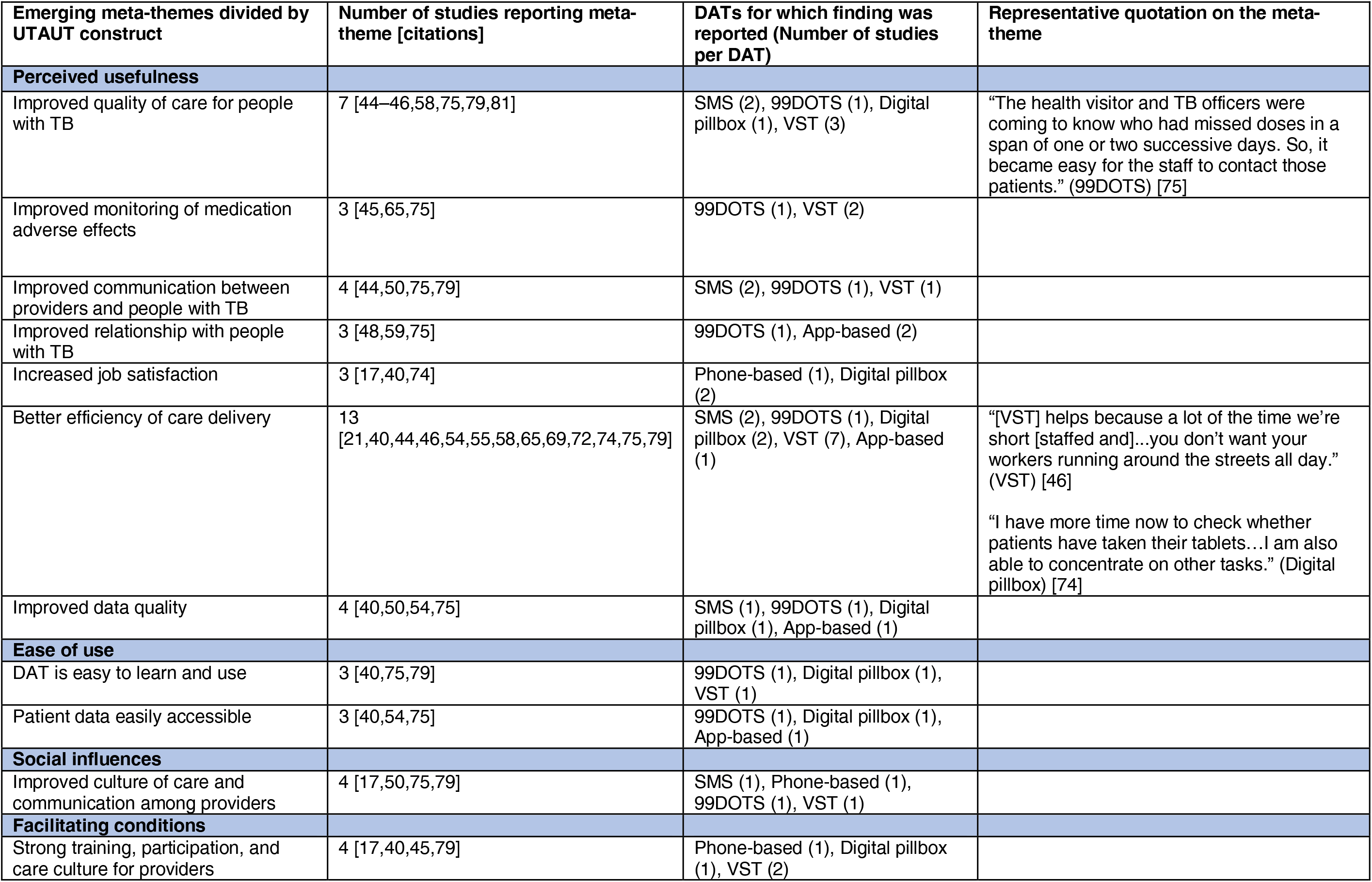
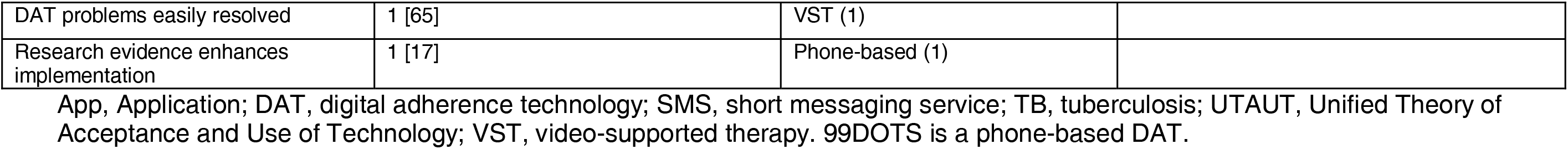
Meta-themes promoting adoption (i.e., DAT uptake by healthcare providers) with findings organized using the UTAUT.

With regard to the UTAUT construct of ease of use, meta-themes that emerged reflected provider appreciation of DATs that were easy to learn and use and that made data from people with TB easily accessible.[40,54,75,79] With regard to the UTAUT construct of social influences, one meta-theme emerged, namely appreciation of DATs that improved the culture of care and communication among healthcare providers.[17,50,75,79] With regard to the UTAUT construct of facilitating conditions, the meta-theme with the most findings pertained to situations in which the DAT was implemented in the context of strong training (e.g., with provision of computer skills or professional development), participation, and care culture for providers (e.g., settings where person-centered care was emphasized).[17,40,45,79]

### Contextual factors limiting DAT adoption by healthcare providers

With regard to the UTAUT construct of perceived usefulness, meta-themes with the most findings related to limiting DAT adoption were negative impact on workload or employment concerns and suboptimal accuracy of adherence data (Table 5; findings informing each meta-theme for adoption are provided in online supplementary appendix 3). DAT interventions sometimes increased the work required of providers (e.g., from time spent training people in DAT use) or raised concerns about job losses due to more efficient care delivery (e.g., fewer providers needed for DOT visits).[17,54,69,73,75,81] Suboptimal accuracy of adherence data referred to challenges experienced in identifying people who were nonadherent to medications, due to inaccuracies in DAT dosing histories.[17,35,45,56,73,77,79] This led to providers contacting many people with TB to verify their dosing histories, or to providers simply presuming that people not reporting doses were taking medications correctly.

**Table 5.** Meta-themes limiting adoption (i.e., DAT use by healthcare providers) with findings organized using the UTAUT.

| Emerging meta-themes divided by UTAUT construct | Number of studies reporting meta-theme [citations] | DATs for which finding was reported (Number of studies per DAT) | Representative quotation on the meta-theme |
| --- | --- | --- | --- |
| **Perceived usefulness** |  |  |  |
| Negative impact on workload or employment concerns | 6 [17,54,69,73,75,81] | Phone-based (1), 99DOTS (2), Digital pillbox (1), VST (1), App-based (1) | "[HIV] drug delivery itself is a big load; we need to give drugs, counsel and also follow-up...[A]dding [99DOTS] has become a burden for us...Because of shortage of manpower, we cannot have a single person constantly looking after TB/HIV patients." (99DOTS) [73] |
| Suboptimal accuracy of adherence data | 7 [17,35,45,56,73,77,79] | Phone-based (1), 99DOTS (1), Digital pillbox (2), VST (3) | Description of a situation faced by a person with TB that led to suboptimal accuracy of dosing information, leading to concerns by healthcare providers:<br>"Since I started going to work, I haven't been using [the digital pillbox] that much because I cannot bring it along. It's very cumbersome...I put [the medications] in my backpack." (Digital pillbox) [35]<br>"Not more than 50% [of patients give a missed call]. But they will take tablet[s]." (99DOTS) [73] |
| Reduced in-person patient-provider interactions negatively impact care | 2 [54,69] | VST (1), App-based (1) |  |
| **Ease of use** |  |  |  |
| Challenges contacting patients | 1 [30] | Phone-based (1) |  |
| **Social influences** |  |  |  |
| Decreased communication among HCPs | 2 [67,73] | 99DOTS (2) |  |
| **Facilitating conditions** |  |  |  |
| Suboptimal DAT function | 7 [44,56,67,69,73–75] | SMS (1), 99DOTS (3), Digital pillbox (1), VST (2) | "It takes 72 hours for the [digital pillbox] dashboard to show that the patient has taken medications. This makes it difficult for us to monitor the patient's drug intake...We cannot take action as promptly..." (Digital pillbox) [74] |
| Loss or destruction of the DAT | 4 [45,51,62,77] | SMS (1), Digital pillbox (2), VST (1) |  |
| Inadequate availability of technology | 1 [40] | Digital pillbox (1) |  |
| Complex cellular accessibility challenges (systemic level) | 6 [17,40,67,69,73,79] | Phone-based (1), 99DOTS (2), Digital pillbox (1), VST |  |
|  |  | (2) |  |
| Insufficient training or technology skills for providers | 5 [17,40,67,73,75] | Phone-based (1), 99DOTS (3), Digital pillbox (1) | "There were few trainings...there was no joint training for both [TB clinic] and [HIV clinic] staff." (99DOTS) [73] |
| Insufficient staff for DAT implementation | 3 [17,73,75] | Phone-based (1), 99DOTS (2) |  |
| Poor management of DAT program | 3 [67,73,75] | 99DOTS (3) |  |
| Suboptimal culture of care | 2 [17,73] | Phone-based (1), 99DOTS (1) |  |
| Government barriers to implementation or sustainment | 3 [17,54,69] | Phone-based (1), VST (1), App-based (1) |  |
| Equity challenges | 1 [69] | VST (1) |  |

App, Application; DAT, digital adherence technology; SMS, short messaging service; TB, tuberculosis; UTAUT, Unified Theory of Acceptance and Use of Technology; VST, video-supported therapy. 99DOTS is a phone-based DAT.

With regard to the UTAUT construct of ease of use, one meta-theme emerged, regarding challenges contacting people with TB, which included findings from only one study.[30] Similarly, for the UTAUT construct of social influences, one meta-theme emerged on decreased communication among healthcare providers, specifically describing challenges coordinating care between TB and HIV programs in India after 99DOTS implementation.[67,73]

With regard to the UTAUT construct of facilitating conditions, the four themes with the most frequent findings were: suboptimal DAT function, loss or destruction of DATs, complexity of cellular accessibility challenges at a systemic level, and insufficient training or technology skills for providers. From the perspective of providers, suboptimal DAT function referred to challenges such as inability to add more than one phone number per patient, inability to add information for treatment supporters, or infrequent updating of digital dosing histories.[44,56,67,69,73–75] Complex cellular accessibility challenges at a systemic level were a major barrier to adoption, with problems including failure of DAT digital records due to power shortages, weak network coverage or Wi-Fi at clinical sites, or problems with video quality or receipt.[17,40,67,69,73,79]

## DISCUSSION

In this paper, we identified numerous contextual factors promoting or limiting the implementation of TB DATs, specifically the reach of these technologies among people with TB and their adoption by healthcare providers. These diverse contextual factors provide insights into reasons for suboptimal engagement of people with TB and healthcare providers as identified in the companion scoping review of DAT implementation outcomes,[8] while also partly explaining the mixed effectiveness of DATs for improving TB clinical outcomes and their variable accuracy for measuring adherence, especially in low- and lower-middle-income country settings.[5,6] By providing insights into what people with TB and healthcare providers value in TB care, this review may also inform strategies for tailoring DAT interventions to enhance future reach, adoption, and effectiveness.

Complex cellular accessibility challenges were a major cross-cutting meta-theme limiting both reach and adoption. These challenges help explain the variability in adoption and serial drop-offs in engagement adversely affecting DAT reach—due to poor cellphone access, lack of initial uptake, and lack of sustained use—that were quantified in the companion DAT implementation outcomes review.[8] Most of these findings were reported from low- and lower-middle-income country settings with less robust cellular network coverage, though these barriers were also experienced in upper- and upper-middle-income country settings. Although optimism around the global expansion of cellular networks has driven use of DATs in TB care, both DAT implementation scoping reviews suggest that the realities of cellular accessibility are more complicated and may undermine DAT intervention programs. Cellular network coverage may be particularly challenging for people with TB, as TB disproportionately affects individuals experiencing socioeconomic disadvantage.[82,83]

Among people with TB, other meta-themes on barriers to ease of use that limited reach—such as literacy and language barriers, technical complexity of some DATs, and suboptimal DAT function—may have contributed to technology fatigue. TB-related stigma was an important meta-theme limiting reach, because use of DATs to support adherence (e.g., via audible or visual reminders) or report dose ingestion often made the act of pill-taking more conspicuous to family or coworkers, especially when compared to self-administered therapy without DAT monitoring. Inequitable access to DATs—by sex, age, education, income, or urban versus rural location—was another major meta-theme limiting reach. Although a recent study suggests that some DAT-based interventions could help reduce inequity in TB treatment outcomes,[84] findings of differential access to DATs in some settings also raises concern that these interventions could increase inequity in outcomes in the overall population of people with TB.

Meta-themes promoting DAT reach also provide general insights into what people value in their TB care. The meta-theme with the most robust findings related to perceptions of improved medication adherence behavior, with several studies reporting that DAT interventions helped people form habits around pill-taking. This finding is perhaps not surprising, given that supporting medication adherence is the main premise of DAT interventions.[1] However, people with TB also valued other benefits of DATs. For example, people with TB appreciated when DATs enhanced their ability to communicate with healthcare providers, provided enhanced access to TB information, improved monitoring of medication adverse effects, and fostered a general sense of being “cared for” by the health system. This last finding resonates with previous research on digital pillboxes among people with HIV in Uganda, in which real-time monitoring was experienced as “being seen” by the health system, thereby motivating people to “take responsibility” for medication adherence.[85]

Our findings suggest that perceptions of DATs by people with TB were strongly shaped by their perceptions of alternative models of TB care. For example, alternative care models sometimes included clinic-based DOT, in which people with TB visit clinics daily for observation of dosing by healthcare providers, which may affect employment and autonomy for people with TB.[86,87] When this care model was the point of reference, people with TB tended to perceive DAT-based care models as resulting in greater convenience and lower stigma. Conversely, if self-administered therapy (i.e., people taking medications on their own at home) was the point of reference, people tended to report more technology fatigue and increased stigma with DAT-based care models. Similarly, a recent systematic review found that the cost-effectiveness of TB DATs was highly influenced by the alternative care model to which the DAT-based model was being compared, with DAT-based models being more likely to be cost-effective when compared with more intensive in-person DOT.[88] As such, findings from DAT studies—whether related to convenience, stigma, or cost-effectiveness—are often as much a reflection of challenges or benefits of prior or alternative care models as they are reflections of the DAT interventions themselves.[89]

With regard to adoption, healthcare providers valued DATs when they improved the efficiency of care, by enabling fewer providers to manage more people with TB in a less time- and resource-intensive manner. Healthcare providers also valued potential improvements in quality of care due to DAT interventions, including benefits related to better identification of medication nonadherence, improved communication with people with TB, and enhanced identification of medication adverse effects. Importantly, one meta-theme indicated that some DAT interventions also improved communication among healthcare providers themselves (i.e., within the care team) in a manner that may have improved the overall culture of care delivery.

A major meta-theme limiting adoption by healthcare providers was negative impacts of DAT-based care models on provider workloads (e.g., due to new needs to counsel people with TB on the DAT) or employment (e.g., fear of losing their jobs due to increased efficiencies in care delivery due to DATs). Healthcare providers were also sometimes frustrated due to the suboptimal accuracy of adherence data (i.e., digital dosing histories) compiled by DATs, consistent with problems with, or variability in, DAT accuracy reported in prior studies and a recent systematic review.[6,90,91] Under-reporting of dose-taking, even when people with TB usually were taking their medications, resulted in providers having to regularly reach out to most people with TB to further assess their medication adherence.[17] In some settings, provider outreach to people with TB dropped off, as providers began to assume that most people who were not reporting doses were probably taking their medications.[73] This finding highlights the importance of evaluating the accuracy of DATs in real world implementation, as inaccurate digital dosing data undermines the confidence of healthcare providers in these interventions.

Strengths of this review include use of a unique methodological approach that systematically synthesized qualitative and quantitative contextual data to identify diverse meta-themes informing reach and adoption of DATs. A limitation of this scoping review is that we did not rate study quality. However, most systematic reviews of qualitative research do not eliminate studies based on quality; instead, qualitative systematic reviews tend to accentuate findings from higher quality studies, as lower-quality studies tend to have less data-rich findings, which we also found to be the case in this review.[9,15] Another limitation of this review is that relatively few studies reported findings on DAT adoption by healthcare providers. This limitation highlights a need for further research on DAT adoption, especially given highly variable DAT adoption by healthcare providers in some settings.[8,92]

## CONCLUSIONS

This scoping review has identified diverse contextual factors that may influence the reach of DATs among people with TB and adoption of these technologies by healthcare providers. These findings may inform selection of appropriate settings for DAT implementation, redesign of DATs to better address the needs of people with TB and healthcare providers, and implementation strategies to improve DAT uptake. A major finding of this review is that, despite increasing access to cellular networks globally, people with TB face diverse challenges to cellular accessibility that may undermine the effectiveness of DAT interventions, especially in low- and middle-income country settings. As such, selection of settings with high cellular accessibility, or efforts to increase cellular accessibility (e.g., provision of cellphones) may be critical to improve the reach and effectiveness of DAT interventions.

While people with TB value potential benefits of DATs for improving medication adherence, they also value other benefits such as enhanced communication with healthcare providers, ability to report medication adverse effects, and use of DAT platforms to gain important information about TB, all of which may foster a sense of being care for by the health system. Similarly, in addition to improvements in the efficiency of care delivery, healthcare providers also value the ability to communicate more easily with people with TB and with other providers, which may improve the overall culture of care. Future DAT-based interventions should move beyond solely focusing on digital observation of dosing and measurement of adherence, by leveraging DATs to facilitate human interactions and address structural barriers to care.[3,93] Focusing on what people with TB and healthcare providers value may enhance the future implementation, effectiveness, and public health impact of these technologies.

## Data Availability

All data produced in the present study are available upon reasonable request to the authors

https://www.medrxiv.org/content/10.1101/2024.01.31.24302115v1.full-text

https://www.medrxiv.org/content/10.1101/2024.06.11.24308660v1

https://www.medrxiv.org/content/10.1101/2024.05.24.24307907v1

## Contributors

RS, SB, KF, and KS conceptualized the study and designed the protocol for the scoping review. MS, SB, CC, CK, MZ, and NF led identification of eligible articles for the review, and RS, KF, and KS resolved conflicts during the article identification process. SB, CC, KF, RS, and NF extracted relevant findings from articles included in the review. SB and RS organized findings and conducted thematic synthesis to identify meta-themes from the findings. BP helps with organization and reporting of findings in the supplementary appendices. SB and RS wrote the first draft of the manuscript. All authors provided critical revisions to the initial manuscript draft and approved the final paper.

## Online supplemental material

Online supplementary appendix 1. PRISMA scoping reviews checklist and search strategy

Online supplementary appendix 2. Findings informing “reach” meta-themes

Online supplementary appendix 3. Findings informing “adoption” meta-themes

## REFERENCES

1 Subbaraman R, de Mondesert L, Musiimenta A, et al. Digital adherence technologies for the management of tuberculosis therapy: mapping the landscape and research priorities. BMJ Glob Health. 2018;3:e001018. doi: 10.1136/bmjgh-2018-001018

2 Fielding K, Subbaraman R, Khan A, et al. The use of digital technologies in adherence to anti-tuberculosis treatment. Digital Respiratory Healthcare (ERS Monograph). Sheffield: European Respiratory Society 2023:170–84.

3 Subbaraman R, Fielding K. Putting technology to the test in tuberculosis care. Lancet. 2024;403:878–9. doi: 10.1016/S0140-6736(24)00412-4

4 Subbaraman R, Fielding K, Thies W, et al. Randomized trial findings suggest an uncertain trail ahead for TB digital adherence technologies. International Journal of Tuberculosis and Lung Disease. 2022;26:378–9.

5 Mohamed MS, Zary M, Kafie C, et al. The impact of digital adherence technologies on health outcomes in tuberculosis: A systematic review and meta-analysis. medRxiv. 2024;2024.01.31.24302115. doi: 10.1101/2024.01.31.24302115

6 Zary M, Mohamed MS, Kafie C, et al. The Performance of Digital Technologies for Measuring Tuberculosis Medication Adherence: A Systematic Review. medRxiv. 2024;2024.05.24.24307886. doi: 10.1101/2024.05.24.24307886

7 Glasgow RE, Vogt TM, Boles SM. Evaluating the public health impact of health promotion interventions: the RE-AIM framework. Am J Public Health. 1999;89:1322–7. doi: 10.2105/ajph.89.9.1322

8 Chilala C, Foster N, Bahukudumbi S, et al. Implementation outcomes of tuberculosis digital adherence technologies: a scoping review using the RE-AIM framework. medRxiv. 2024;2024.06.11.24308660. doi: 10.1101/2024.06.11.24308660

9 Lachal J, Revah-Levy A, Orri M, et al. Metasynthesis: An Original Method to Synthesize Qualitative Literature in Psychiatry. Front Psychiatry. 2017;8:269. doi: 10.3389/fpsyt.2017.00269

10 Venkatesh V, Morris M, Davis G, et al. User Acceptance of Information Technology: Toward a Unified View. MIS Quarterly. 2003;27:425–78. doi: 10.2307/30036540

11 Tricco AC, Lillie E, Zarin W, et al. PRISMA Extension for Scoping Reviews (PRISMA-ScR): Checklist and Explanation. Ann Intern Med. 2018;169:467–73. doi: 10.7326/M18-0850

12 Pai M, McCulloch M, Gorman JD, et al. Systematic reviews and meta-analyses: an illustrated, step-by-step guide. Natl Med J India. 2004;17:86–95.

13 Noblit G, Hare R. Meta-ethnography: Synthesizing qualitative studies. Newbury Park, USA: Sage 1988.

14 Noyes J, Booth A, Cargo M, et al. Chapter 21: Qualitative evidence. Cochrane Handbook for Systematic Reviews of Interventions. Cochrane 2023.

15 Munro SA, Lewin SA, Smith HJ, et al. Patient adherence to tuberculosis treatment: a systematic review of qualitative research. PLoS Med. 2007;4:e238. doi: 10.1371/journal.pmed.0040238

16 Cross A, Gupta N, Liu B, et al. 99DOTS: A Low-Cost Approach to Monitoring and Improving Medication Adherence. Ahmedabad, India: Assoc Computing Machinery 2019.

17 Bardosh KL, Murray M, Khaemba AM, et al. Operationalizing mHealth to improve patient care: A qualitative implementation science evaluation of the WelTel texting intervention in Canada and Kenya. Globalization and Health. 2017;13:87. doi: 10.1186/s12992-017-0311-z

18 Bassett IV, Coleman SM, Giddy J, et al. Sizanani: a Randomized Trial of Health System Navigators to Improve Linkage to HIV and TB Care in South Africa. J Acquir Immune Defic Syndr. 2016;73:154–160. doi: 10.1097/QAI.0000000000001025

19 Bediang G, Stoll B, Elia N, et al. SMS reminders to improve adherence and cure of tuberculosis patients in Cameroon (TB-SMS Cameroon): a randomised controlled trial. BMC Public Health. 2018;18:583–583.

20 Belknap R, Weis S, Brookens A, et al. Feasibility of an Ingestible Sensor-Based System for Monitoring Adherence to Tuberculosis Therapy. PLoS ONE. 2013;8:e53373.

21 Bendiksen R, Ovesen T, Asfeldt AM, et al. Use of video call in the treatment of tuberculosis disease in Northern Norway. Tidsskrift for Den Norske Laegeforening. 2020;140:35–40. doi: 10.4045/tidsskr.19.0322

22 Bionghi N, Daftary A, Maharaj B, et al. Pilot evaluation of a second-generation electronic pill box for adherence to Bedaquiline and antiretroviral therapy in drug-resistant TB/HIV co-infected patients in KwaZulu-Natal, South Africa. BMC Infectious Diseases. 2018;18:171.

23 Bommakanti KK, Smith LL, Liu L, et al. Requiring smartphone ownership for mHealth interventions: who could be left out? BMC Public Health. 2020;20:81.

24 Browne SH, Umlauf A, Tucker AJ, et al. Wirelessly observed therapy compared to directly observed therapy to confirm and support tuberculosis treatment adherence: A randomized controlled trial. PLoS Medicine. 2019;16:e1002891.

25 Buchman T, Cabello C. A New Method to Directly Observe Tuberculosis Treatment: Skype Observed Therapy, a Patient-Centered Approach. Journal of Public Health Management and Practice. 2017;23:175–7.

26 Burzynski J, Mangan JM, Lam CK, et al. In-Person vs Electronic Directly Observed Therapy for Tuberculosis Treatment Adherence: A Randomized Noninferiority Trial. JAMA Network Open. 2022;5:e2144210.

27 Chen SH, Wang I, Hsu HL, et al. Advantage in privacy protection by using synchronous video observed treatment enhances treatment adherence among patients with latent tuberculosis infection. Journal of Infection and Public Health. 2020;13:1354–9.

28 Chuck C, Robinson E, Macaraig M, et al. Enhancing management of tuberculosis treatment with video directly observed therapy in New York City. International Journal of Tuberculosis and Lung Disease. 2016;20:588–93.

29 Cox SN, Elf J, Lokhande R, et al. Mobile phone access and comfort: Implications for HIV and tuberculosis care in India and South Africa. Open Forum Infectious Diseases. 2018;5:S162.

30 Daftary A, Hirsch-Moverman Y, Kassie G, et al. A Qualitative Evaluation of the Acceptability of an Interactive Voice Response System to Enhance Adherence to Isoniazid Preventive Therapy Among People Living with HIV in Ethiopia. AIDS & Behavior. 2017;21:3057–67.

31 Das Gupta D, Patel A, Saxena D, et al. Choice-Based Reminder Cues: Findings From an mHealth Study to Improve Tuberculosis (TB) Treatment Adherence Among the Urban Poor in India. World Medical and Health Policy. 2020;12:163–81.

32 de Sumari-de Boer IM, van den Boogaard J, Ngowi KM, et al. Feasibility of Real Time Medication Monitoring Among HIV Infected and TB Patients in a Resource-Limited Setting. AIDS and Behavior. 2016;20:1097–107. doi: 10.1007/s10461-015-1254-0

33 DeMaio J, Schwartz L, Cooley P, et al. The application of telemedicine technology to a directly observed therapy program for tuberculosis: A pilot project. Clinical Infectious Diseases. 2001;33:2082–4. doi: 10.1086/324506

34 Dessie Gashu K, Nurhussien F, Mamuye A, et al. Developing and Piloting TB Medication and Refilling Reminder System in Ethiopia. Studies in health technology and informatics. 2020;270:1251–2. doi: 10.3233/SHTI200387

35 Drabarek D, Anh NT, Nhung NV, et al. Implementation of Medication Event Reminder Monitors among patients diagnosed with drug susceptible tuberculosis in rural Viet Nam: a qualitative study. PLoS ONE. 2019;14:e0219891. doi: 10.1371/journal.pone.0219891

36 Garfein RS, Liu L, Cuevas-Mota J, et al. Evaluation of recorded video-observed therapy for anti-tuberculosis treatment. International Journal of Tuberculosis and Lung Disease. 2020;24:520–5.

37 Garfein RS, Collins K, Munoz F, et al. Feasibility of tuberculosis treatment monitoring by video directly observed therapy: A binational pilot study. International Journal of Tuberculosis and Lung Disease. 2015;19:1057–64.

38 Gashu KD, Gelaye KA, Tilahun B. Feasibility, acceptability and challenges of phone reminder system implementation for tuberculosis pill refilling and medication in Northwest Ethiopia. Research Square. 2021;Version 1. doi: 10.21203/rs.3.rs-229284/v1

39 Gassanov MA, Feldman LJ, Sebastian A, et al. The use of videophone for directly observed therapy for the treatment of tuberculosis. Canadian Journal of Public Health. 2013;104:e272.

40 Getachew E, Woldeamanuel Y, Manyazewal T. Digital health interventions in the clinical care and treatment of tuberculosis and hiv in central Ethiopia: An initial provider perceptions and acceptability study using the unified theory of acceptance and use of technology model. Int J Mycobacteriol. 2022;11:1–9.

41 Guo X, Yang Y, Takiff HE, et al. A Comprehensive App That Improves Tuberculosis Treatment Management Through Video-Observed Therapy: Usability Study. JMIR mHealth and uHealth. 2020;8:e17658.

42 Guo P, Qiao W, Sun Y, et al. Telemedicine Technologies and Tuberculosis Management: a Randomized Controlled Trial. Telemedicine journal and e-health. 2020;26:1150–1156.

43 Hermans SM, Elbireer S, Tibakabikoba H, et al. Text messaging to decrease tuberculosis treatment attrition in TB-HIV coinfection in Uganda. Patient Preference and Adherence. 2017;11:1479–87. doi: 10.2147/PPA.S135540

44 Hirsch-Moverman Y, Daftary A, Yuengling KA, et al. Using mhealth for HIV/TB treatment support in lesotho: Enhancing patient-provider communication in the start study. Journal of Acquired Immune Deficiency Syndromes. 2017;74:S37–43.

45 Hoffman JA, Cunningham JR, Suleh AJ, et al. Mobile Direct Observation Treatment for Tuberculosis Patients. A Technical Feasibility Pilot Using Mobile Phones in Nairobi, Kenya. American Journal of Preventive Medicine. 2010;39:78–80.

46 Holzman SB, Zenilman A, Shah M. Advancing patient-centered care in tuberculosis management: A mixed-methods appraisal of video directly observed therapy. Open Forum Infectious Diseases. 2018;5:ofy046. doi: 10.1093/ofid/ofy046

47 Holzman SB, Atre S, Sahasrabudhe T, et al. Use of Smartphone-Based Video Directly Observed Therapy (vDOT) in Tuberculosis Care: Single-Arm, Prospective Feasibility Study. JMIR Form Res. 2019;3:e13411. doi: 10.2196/13411

48 Horter S, Stringer B, Venis S, et al. ‘I can also serve as an inspiration’: A qualitative study of the TB&Me blogging experience and its role in MDRTB treatment. PLoS ONE. 2014;9:e108591.

49 Iribarren SJ, Rodriguez Y, Lin L, et al. Converting and expanding a mobile support intervention: Focus group and field-testing findings from individuals in active tuberculosis treatment. International Journal of Medical Informatics. 2020;136:104057.

50 Iribarren SJ, Sward KA, Beck SL, et al. Qualitative evaluation of a text messaging intervention to support patients with active tuberculosis: implementation considerations. JMIR Mhealth Uhealth. 2015;3:e21. doi: 10.2196/mhealth.3971

51 Iribarren S, Beck S, Pearce PF, et al. TextTB: A Mixed Method Pilot Study Evaluating Acceptance, Feasibility, and Exploring Initial Efficacy of a Text Messaging Intervention to Support TB Treatment Adherence. Tuberc Res Treat. 2013;2013:349394. doi: 10.1155/2013/349394

52 Kalita J, Pandey PC, Shukla R, et al. Feasibility and usefulness of tele-follow-up in the patients with tuberculous meningitis. Transactions of the Royal Society of Tropical Medicine and Hygiene. 2021;115:1153–9. doi: 10.1093/trstmh/trab06

53 Khachadourian V, Truzyan N, Harutyunyan A, et al. People-centred care versus clinic-based DOT for continuation phase TB treatment in Armenia: a cluster randomized trial. BMC Pulm Med. 2020;20:105. doi: 10.1186/s12890-020-1141-y

54 Kopanitsa G. A Qualitative Study of the Barriers and Opportunities for Adoption of Web-Portals for Doctors and Patients in Russia. Journal of Medical Systems. 2017;41:62. doi: 10.1007/s10916-017-0713-8

55 Krueger K, Ruby D, Cooley P, et al. Videophone utilization as an alternative to directly observed therapy for tuberculosis. International Journal of Tuberculosis and Lung Disease. 2010;14:779–81. doi: 10.5588/ijtld.10.0210

56 Lam CK, McGinnis Pilote K, Haque A, et al. Using Video Technology to Increase Treatment Completion for Patients With Latent Tuberculosis Infection on 3-Month Isoniazid and Rifapentine: An Implementation Study. J Med Internet Res. 2018;20:e287. doi: 10.2196/jmir.9825

57 Liu X, Lewis JJ, Zhang H, et al. Effectiveness of Electronic Reminders to Improve Medication Adherence in Tuberculosis Patients: A Cluster-Randomised Trial. PLOS Med. 2015;12:e1001876. doi: 10.1371/journal.pmed.1001876

58 Mahmud N, Rodriguez J, Nesbit J. A text message-based intervention to bridge the healthcare communication gap in the rural developing world. Technol Health Care. 2010;18:137–44. doi: 10.3233/THC-2010-0576

59 Milligan H, Iribarren SJ, Chirico C, et al. Insights from participant engagement with the tuberculosis treatment support tools intervention: Thematic analysis of interactive messages to guide refinement to better meet end user needs. International Journal of Medical Informatics. 2021;149:104421. doi: 10.1016/j.ijmedinf.2021.104421

60 Mohammed S, Glennerster R, Khan AJ. Impact of a Daily SMS Medication Reminder System on Tuberculosis Treatment Outcomes: a Randomized Controlled Trial. PLoS ONE. 2016;11:e0162944. doi: 10.1371/journal.pone.0162944

61 Mohammed S, Siddiqi O, Ali O, et al. User engagement with and attitudes towards an interactive SMS reminder system for patients with tuberculosis. Journal of Telemedicine and Telecare. 2012;18:404–8. doi: 10.1258/jtt.2012.120311

62 Moulding TS, Caymittes M. Managing medication compliance of tuberculosis patients in Haiti with medication monitors. International journal of tuberculosis and lung disease. 2002;6:313–319. doi: 10.5588/ijtld.02.0377

63 Navin K, Vadivu G, Maharaj A, et al. A Mobile Health Intervention to Support TB Eradication Programme for Adherence to Treatment and a Novel QR Code Based Technique to Monitor Patient-DOTS Provider Interaction. Advanced Computational and Communication Paradigms. Singapore: Springer 2018:41–54.

64 Nhavoto JA, Grönlund Å, Klein GO. Mobile health treatment support intervention for HIV and tuberculosis in Mozambique: perspectives of patients and healthcare workers. PloS ONE. 2017;12:e0176051. doi: 10.1371/journal.pone.0176051

65 Olano-Soler H, Thomas D, Joglar O, et al. Notes from the Field: Use of Asynchronous Video Directly Observed Therapy for Treatment of Tuberculosis and Latent Tuberculosis Infection in a Long-Term-Care Facility - Puerto Rico, 2016-2017. MMWR Morbidity and Mortality Weekly Report. 2017;66:1386–7. doi: 10.15585/mmwr.mm6650a5

66 Person AK, Blain ML, Jiang H, et al. Text messaging for enhancement of testing and treatment for tuberculosis, human immunodeficiency virus, and syphilis: a survey of attitudes toward cellular phones and healthcare. Telemed J E Health. 2011;17:189–95. doi: 10.1089/tmj.2010.0164

67 Prabhu A, Agarwal U, Singla N, et al. ‘99DOTS’ techno-supervision for tuberculosis treatment - a boon or a bane? Exploring challenges in its implementation at a tertiary centre in Delhi, India. Indian J Tuberc. 2020;67:46–53. doi: 10.1016/j.ijtb.2019.08.010

68 Ratchakit-Nedsuwan R, Nedsuwan S, Sawadna V, et al. Ensuring tuberculosis treatment adherence with a mobile-based CARE-call system in Thailand: a pilot study. Infect Dis (Lond). 2020;52:121–129. doi: 10.1080/23744235.2019.1688862

69 Ritter LA, Mei Wa K, Nasseri, et al. California Public Health Departments Remotely Treat Tuberculosis: Outcomes & Opportunities. Californian Journal of Health Promotion. 2017;15:37–45.

70 Sinkou H, Hurevich H, Rusovich V, et al. Video-observed treatment for tuberculosis patients in Belarus: Findings from the first programmatic experience. European Respiratory Journal. 2017;49:1602049.

71 Stagg HR, Lewis JJ, Liu X, et al. Temporal Factors and Missed Doses of Tuberculosis Treatment. A Causal Associations Approach to Analyses of Digital Adherence Data. Annals of the American Thoracic Society. 2020;17:438–449. doi: 10.1513/AnnalsATS.201905-394OC

72 Story A, Aldridge RW, Smith CM, et al. Smartphone-enabled video-observed versus directly observed treatment for tuberculosis: a multicentre, analyst-blinded, randomised, controlled superiority trial. The Lancet. 2019;393:1216–24. doi: 10.1016/S0140-6736(18)32993-3

73 Thekkur P, Kumar AMV, Chinnakali P, et al. Outcomes and implementation challenges of using daily treatment regimens with an innovative adherence support tool among HIV-infected tuberculosis patients in Karnataka, India: a mixed-methods study. Global Health Action. 2019;12:1–11. doi: 10.1080/16549716.2019.1568826

74 Thomas BE, Kumar JV, Periyasamy M, et al. Acceptability of the Medication Event Reminder Monitor for Promoting Adherence to Multidrug-Resistant Tuberculosis Therapy in Two Indian Cities: Qualitative Study of Patients and Health Care Providers. Journal of Medical Internet Research. 2021;23:e23294.

75 Thomas BE, Kumar JV, Onongaya C, et al. Explaining Differences in the Acceptability of 99DOTS, a Cell Phone-Based Strategy for Monitoring Adherence to Tuberculosis Medications: Qualitative Study of Patients and Health Care Providers. JMIR Mhealth Uhealth. 2020;8:e16634.

76 Ting NCH, El-Turk N, Chou MSH, et al. Patient-perceived treatment burden of tuberculosis treatment. PLoS ONE. 2020;15:e0241124.

77 Trajman A, Long R, Zylberberg D, et al. Factors associated with treatment adherence in a randomised trial of latent tuberculosis infection treatment. International Journal of Tuberculosis and Lung Disease. 2010;14:551–559.

78 van den Boogaard J, Lyimo RA, Boeree MJ, et al. Electronic monitoring of treatment adherence and validation of alternative adherence measures in tuberculosis patients: A pilot study. Bulletin of the World Health Organization. 2011;89:632–9.

79 Wade VA, Karnon J, Eliott JA, et al. Home Videophones Improve Direct Observation in Tuberculosis Treatment: A Mixed Methods Evaluation. PLoS ONE. 2012;7:e50155. doi: 10.1371/journal.pone.0050155

80 Wade V, Izzo J, Hamlyn J. Videophone delivery of Medication Management in Community Nursing. Electronic Journal of Health Informatics. 2009;4:5. doi: 10.1007/8754_2011_26

81 Wang N, Zhang H, Zhou Y, et al. Using electronic medication monitoring to guide differential management of tuberculosis patients at the community level in China. BMC Infectious Diseases. 2019;19:844. doi: 10.1186/s12879-019-4521-2

82 Oxlade O, Murray M. Tuberculosis and poverty: why are the poor at greater risk in India? PLoS ONE. 2012;7:e47533. doi: 10.1371/journal.pone.0047533

83 Jhaveri TA, Jhaveri D, Galivanche A, et al. Barriers to engagement in the care cascade for tuberculosis disease in India: A systematic review of quantitative studies. PLoS Med. 2024;21:e1004409. doi: 10.1371/journal.pmed.1004409

84 Boutilier JJ, Yoeli E, Rathauser J, et al. Can digital adherence technologies reduce inequity in tuberculosis treatment success? Evidence from a randomised controlled trial. BMJ Global Health. 2022;7:e010512. doi: 10.1136/bmjgh-2022-010512

85 Ware NC, Pisarski EE, Tam M, et al. The Meanings in the messages: how SMS reminders and real-time adherence monitoring improve antiretroviral therapy adherence in rural Uganda. AIDS. 2016;30:1287–94. doi: 10.1097/QAD.0000000000001035

86 Yellappa V, Lefèvre P, Battaglioli T, et al. Coping with tuberculosis and directly observed treatment: a qualitative study among patients from South India. BMC Health Serv Res. 2016;16:283. doi: 10.1186/s12913-016-1545-9

87 Sagbakken M, Frich JC, Bjune GA, et al. Ethical aspects of directly observed treatment for tuberculosis: a cross-cultural comparison. BMC Med Ethics. 2013;14:25. doi: 10.1186/1472-6939-14-25

88 Kafie C, Mohamed MS, Zary M, et al. Cost and Cost-effectiveness of Digital Adherence Technologies for Support of Treatment for Tuberculosis: A systematic review. medRxiv. 2024;2024.05.24.24307907. doi: 10.1101/2024.05.24.24307907

89 Subbaraman R, Haberer JE, Fielding K. Intention to Treat or per Protocol? Overly Optimistic Findings Regarding the Cost-Effectiveness of 99DOTS, a Tuberculosis Digital Adherence Technology. Value in healthfI: the journal of the International Society for Pharmacoeconomics and Outcomes Research. Published Online First: 2022. doi: 10.1016/j.jval.2022.09.008

90 Thomas BE, Kumar JV, Chiranjeevi M, et al. Evaluation of the Accuracy of 99DOTS, a Novel Cellphone-based Strategy for Monitoring Adherence to Tuberculosis Medications: Comparison of DigitalAdherence Data with Urine Isoniazid Testing. Clinical Infectious Diseases. 2020;71:E513–6. doi: 10.1093/cid/ciaa333

91 Subbaraman R, Thomas BE, Kumar JV, et al. Measuring Tuberculosis Medication Adherence: A Comparison of Multiple Approaches in Relation to Urine Isoniazid Metabolite Testing Within a Cohort Study in India. Open Forum Infect Dis. 2021;8:ofab532.

92 Chen A, Kumar R, Baria RK, et al. Impact of the 99DOTS digital adherence technology on tuberculosis treatment outcomes in North India: a pre-post study. BMC Infectious Diseases. 2023;23:504. doi: 10.1186/s12879-023-08418-2

93 Subbaraman R, Thomas BE, Kumar JV, et al. Understanding Nonadherence to Tuberculosis Medications in India Using Urine Drug Metabolite Testing: A Cohort Study. Open Forum Infect Dis. 2021;8:ofab190. doi: 10.1093/ofid/ofab190

