## Supplemental Table 1 for "Contextual factors influencing implementation of tuberculosis digital adherence technologies: a scoping review guided by the RE-AIM framework": Supplementary Appendix 2 - Findings informing adoption meta-themes.docx

Tufts University School of Medicine

### Adoption (Positive contextual factors)

#### Perceived usefulness—Positive findings

*Table 1: Improves monitoring of medication adverse effects.*

| Author Year | Findings |
| --- | --- |
| Video supported therapy | |
| Olano-Soler 2017 [1] | Medical director observed patients over asynchronous videos for signs of hepatoxicity. |
| Hoffman 2010 [2] | Allows for proactive and real time intervention on adverse effects and provision of counseling. |
| 99DOTS | |
| Thomas 2020 [3] | 99DOTS strengthened the patient-HCP relationship and resulted in early identification of medication adverse effects: “*There was one patient who was registered for 99DOTS, but she was not calling. When she was contacted... she said she was experiencing some side effects of the drug... So definitely it is helpful.”* (Quotation from a a higher-level administrative officer from Mumbai) |

*Table 2: Improving quality of care for patients.*

| Author Year | Findings |
| --- | --- |
| Short messaging service | |
| Mahmud 2010 [4] | Of short messaging service text communications sent by healthcare providers, 107/1330 (8%) were to request help, 59/1330 (4%) were to request supplies, and 199/1330 (15%) were to report patient symptoms. |
| Hirsch-Moverman 2017 [5] | Voluntary health workers (VHWs) were also more easily able to track and follow-up with patients at risk for nonadherence or treatment interruption. |
| 99DOTS | |
| Thomas 2020 [6] | Perceived improvements by healthcare providers in the quality of care. |

| Digital pillbox | |
| --- | --- |
| Wang 2019 [7] | The electronic medication monitor (EMM) was useful to physicians for improving care: stated by 9/9 (100%) providers (nurses and physicians). |

| Video supported therapy | |
| --- | --- |
| Holzman 2018 [8] | Video supported therapy allows for observed therapy 7 times per week. [This was more than was possible with in-person DOT and was considered an improvement in quality of care] |
| Holzman 2018 [8] | Video supported therapy is effective for monitoring patient adherence: 15/16 (94%). |
| Wade 2012 [9] | More patients were referred to community nursing program for direct observation. [This was considered to be a triage function that improves quality of care] |
| Hoffman 2010 [2] | Provides "a higher level of care" to patients. |
| Hoffman 2010 [2] | Allows for proactive and real time intervention on adverse effects and provision of counseling. |

*Table 3: Improved communication between healthcare providers and patients*

| Author Year | Findings |
| --- | --- |
| Short messaging service | |
| Iribarren 2015 [10] | Providers felt TextTB was particularly helpful for communicating with, and supporting, TB patients in rural areas who were taking self-administered therapy. |
| Hirsch-Moverman 2017 [5] | Some voluntary health workers (VHWs) would call patients nearly every day to remind them to take daily doses and to ensure that they made it to their next clinic visit with the appropriate tools in hand such as sputum samples. This helped to avoid delays at the facility, capture and resolve potential reasons for missing a visit, and prevent nonadherence. |
| 99DOTS | |
| Thomas 2020 [3] | Healthcare providers felt that 99DOTS enhanced the frequency and ease of outreach to patients by phone: “*Actually, due to the 99DOTS program, communication [with the patient] has increased... [99DOTS] is useful...we can call the patient and inform them to take pills, which is beneficial*.” (Quotation from a TB Officer from Mumbai) |
| Video supported therapy | |
| Wade 2012 [9] | Absent patients could be readily called back repeatedly. |

*Table 4: Improved relationship with patients*

| Author Year | Findings |
| --- | --- |
| 99DOTS | |
| Thomas 2020 [3] | Perceived improvements in the patient-healthcare provider relationship. |
| App-based | |
| Milligan 2021 [11] | Healthcare providers felt they established a better relationship with patients. The treatment supporter emphasized the importance of building a trusting relationship with the participants and stated feeling, “*more like a treatment partner than a health professional conducting adherence surveillance*.” |
| Horter 2014 [12] | Enhanced patient-provider relationship because provider read the blog entries to get insights into patients (provides a "human face" to the patient). |

*Table 5: Increased job satisfaction*

| Author Year | Findings |
| --- | --- |
| Phone-based | |
| Bardosh 2017 [13] | Improved work satisfaction for providers due to better connections with patients. |
| Digital pillbox | |
| Thomas 2021 [14] | Some HCPs reported that the decreased workload allowed them to focus more on each patient interaction, as well as other tasks, which increased job satisfaction: *“I have more time now to check whether patients have taken their tablets or not. I am also able to concentrate on other tasks as well, which gives me more satisfaction in my work.”* (Quotation from a health visitor [a local healthcare worker]) |
| Getachew 2022 [15] | Increased job satisfaction. |

*Table 6: Increased efficiency of care delivery*

| Author Year | Findings |
| --- | --- |
| Short messaging service | |
| Mahmud 2010 [4] | 410/1330 (31%) of messages facilitated referral or reporting of patient issues, which saved providers time. 410/1330 |
| Mahmud 2010 [4] | Technology saved providers time by helping to communicate with them about notification of patient death: 75/1330 (6%) were to report a patient death. |
| Mahmud 2010 [4] | Technology saved providers time by helping to communicate with them about notifying patients about missed appointments. |
| Mahmud 2010 [4] | Technology saved providers time by helping to communicate with them about reporting TB medication adherence. |
| Mahmud 2010 [4] | Technology saved providers time by helping to communicate with them about reporting patient questions to doctors. |
| Mahmud 2010 [4] | Technology saved providers time by helping to communicate with them about making requests for acute care. |
| Mahmud 2010 [4] | Technology saved providers time by helping to communicate with them about getting information on appropriate medication dosing. |
| Mahmud 2010 [4] | HBC nurses and the TB coordinator reported $1000 to $2000 in fuel savings, while all short messaging service messages only costed $250 (net savings were $2,750). 648 hours of transportation time were saved for the TB program. 2,048 total workers hours were saved. TB coordinator was able to see an additional 100 TB patients per week. |
| Hirsch-Moverman 2017 [5] | Community-based voluntary health workers (VHWs) appreciated being able to monitor patients over phone, as they faced substantial geographic barriers and unpredictable weather when following patients living in disparate, remote rural areas. |
| 99DOTS | |
| Thomas 2020 [6] | Perceived improvements in the efficiency of care |
| Digital pillbox | |
| Thomas 2021 [14] | Some HCPs reported that the decreased workload allowed them to focus more on each patient interaction, as well as other tasks, which increased job satisfaction: “*I have more time now to check whether patients have taken their tablets or not. I am also able to concentrate on other tasks as well, which gives me more satisfaction in my work.”* (Quotation from a health visitor [local TB healthcare provider]) |
| Getachew 2022 [15] | Reduced workload. |
| Video supported therapy | |
| Holzman 2018 [8] | Video supported therapy was convenient for staff. |
| Holzman 2018 [8] | Video supported therapy may increase clinic capacity (saves provider time, allows for redistribution of tasks). |
| Wade 2012 [9] | Absent patients could be readily called back repeatedly. [Implication was that this saves time for providers rather than conducting home visits] |
| Wade 2012 [9] | Patients who had difficulty taking all their tablets at once could be called in stages. |
| Wade 2012 [9] | Many more patients could be seen in a shift than with a drive-around service. |
| Wade 2012 [9] | The Chest Clinic encouraged other hospitals to also refer to the videophone service. |
| Wade 2012 [9] | More patients were referred to community nursing program for direct observation. |
| Olano-Soler 2017 [1] | Time saved due to asynchronous video DOT included 240 hours of DOT-related activities across all patients, including 1.5 hours of travel per day to the facility and 1.5 hours of observation. Time saved was about 25% of the workload of a healthcare worker. |
| Ritter 2017 [16] | Video supported therapy helps reduce clinic crowding. |
| Ritter 2017 [16] | Helps staff in rural counties where travel times are long. |
| Ritter 2017 [16] | Reduces the number of staff needed, which program managers felt is cost-effective. |
| Ritter 2017 [16] | Very helpful in countries bordering the U.S.-Mexico border, as patients may travel across the border and can still be monitored with video supported therapy. |
| Story 2019 [17] | Average staff time per dose observed was 56 min (SD 54) for community-based DOT (including travel time); 15 min (12) for clinic-based DOT, and 3·2 min (0·5) for video supported therapy. |
| Bendiksen 2020 [18] | A home care nurse reported that the home care service did not have to drive all the way to the patient, and this saved a lot of time. The patients had more freedom to live their lives, and this was a really positive thing. |
| Krueger 2010 [19] | Average savings for staff of driving for in-person DOT was 1818 miles per patient or $684 dollars of money per patient or 3151 minutes of travel time per patient. There was also $1764 staff salary saved per patient to the program. The total average cost saved to the program per patient was $2448. |
| App-based | |
| Kopanitsa 2017 [20] | They had an intention to use the portal instead of traditional paperwork and to save time visiting a patient. |

*Table 7: Improved data quality*

| Author Year | Findings |
| --- | --- |
| Short messaging service | |
| Iribarren 2015 [10] | TextTB helped to identify patient transfer of care. |
| 99DOTS | |
| Thomas 2020 [3] | HCPs generally found the 99DOTS adherence dashboard to be easy to use and preferred electronic data entry into the 99DOTS dashboard, as compared with written records (Q32). They also found it easy to generate reports through the 99DOTS dashboard in a manner that prioritizes high-risk patients, as described in the following quotation: “*It’s easy when it comes to reports. Immediately the report can be pulled out to determine which patients need to be given priority.”* (Quotation from a medical officer [TB physician] from Tamil Nadu) |
| Digital pillbox | |
| Getachew 2022 [15] | Fewer problems with missing data. |
| App-based | |
| Kopanitsa 2017 [20] | Data stored electronically it was unlikely to get lost in comparison with the paper records (lost by the clinics on a regular basis). |

#### Ease of use—Positive findings

*Table 8: The DAT is easy to learn and use*

| Author Year | Findings |
| --- | --- |
| 99DOTS | |
| Thomas 2020 [6] | Dashboard was easy to use. |
| Digital Pillbox | |
| Getachew 2022 [15] | Technology easy to use; training and exposure to the technology makes it easy to use. |
| Video supported therapy | |
| Wade 2012 [9] | The service could be initiated rapidly, without technical support. |

*Table 9: Patient and other data is more easily accessible*

| Author Year | Findings |
| --- | --- |
| 99DOTS | |
| Thomas 2020 [6] | Dashboard was easy to use. |
| Digital Pillbox | |
| Getachew 2022 [15] | Retrieving patient data from electronically recorded documents is easy. |
| App-based | |
| Kopanitsa 2017 [20] | Positive comments on technical aspects of the platform. Information visualization and reporting services of the portal allowed them to have all relevant data on the screen and provide very efficient searching facilities. |

#### Social influences—Positive findings

*Table 10: Improved culture of care and communication among healthcare providers*

| Author Year | Findings |
| --- | --- |
| Short messaging service | |
| Iribarren 2015 [10] | Responses to patient queries sent via short messaging service required collaboration among providers, enhancing collaborative care. |
| Phone-based | |
| Bardosh 2017 [21] | Improved "culture of care" including services, standards, and accountability. |
| 99DOTS | |
| Thomas 2020 [6] | Better coordination and communication among HCPs. |
| Video supported therapy | |
| Wade 2012 [9] | Increased communication about patients occurred. |

#### Facilitating conditions—Positive findings

*Table 11: Strong training, participation, and culture of care enhance adoption*

| Author Year | Findings |
| --- | --- |

| Phone Based | |
| --- | --- |
| Bardosh 2017 [21] | Interactions between providers and patients were more frequent in Canada due to more patient-centered care approaches. |
| Digital pillbox | |
| Getachew 2022 [15] | 43.3% of HCP had a favorable working environment to use the technology. |

| Video supported therapy | |
| --- | --- |
| Wade 2012 [9] | The Chest Clinic initiated education of call center nurses. |
| Hoffman 2010 [2] | Computer trainings and professional development was provided and valued by providers. |
| Wade 2012 [9] | Joint protocol development was undertaken. |

*Table 12: DAT problems easily resolved*

| Author Year | Findings |
| --- | --- |
| Video supported therapy | |
| Olano-Soler 2017 [1] | Asynchronous video DOT facilitated observation successfully despite a poor internet connection, because it did not rely on a consistent connection and did not rely on a cellular plan. |

*Table 13: Research evidence enhances implementation*

| Author Year | Findings |
| --- | --- |
| Phone-based | |
| Bardosh 2017 [21] | Data and research on WelTel provided legitimacy for scale-up and maintenance. |

### Adoption (Negative contextual factors)

#### Perceived usefulness—Negative findings

*Table 14: Increased workload for healthcare providers or employment concerns*

| Author Year | Findings | |
| --- | --- | --- |
| Phone-based | | |
| Bardosh 2017 [21] | 30% to 80% of people in Kenya required callback due to not responding to the short messaging service, which was beyond capacity of providers. | |
| 99DOTS | | |
| Thomas 2020 [3] | Increase in workload with 99DOTS implementation without changes in healthcare provider staffing. | |
| Thekkur 2019 [22] | Antiretroviral therapy clinic staff felt their workload had increased and National TB Programme staff involvement was minimal (corroborated by TB staff). | |
| Digital Pillbox | | |
| Wang 2019 [7] | Workload increased while using the electronic medication monitor (EMM): for 8/9 providers (physicians and nurses). | |
| Wang 2019 [7] | Increased workload was moderate while using the electronic medication monitor (EMM): for 7/9 providers (physicians and nurses). | |
| Video supported therapy | | |
| Ritter 2017 [16] | | Healthcare workers were concerned about losing their jobs (though they were reassured). |
| App-based | | |
| Kopanitsa 2017 [20] | That they expected that the tool would improve their communication with patients, but would not result in any additional work for them to do (duplicate their recommendations; unnecessary requests from patients). | |

*Table 15: Suboptimal accuracy of adherence data*

| Author Year | | Findings |
| --- | --- | --- |
| Phone Based | | |
| Bardosh 2017 [21] | | 30% to 80% of people in Kenya required callback due to not responding to the short messaging service, which was beyond capacity of providers. |
| 99DOTS | | |
| Thekkur 2019 [22] | | Low proportion of patients calling 99DOTS led providers to presume patients are taking their tablets and not take actions for non-reported doses. |
| Thekkur 2019 [22] | | Healthcare providers started believing that ‘not getting missed call’ is because of these challenges rather than ‘not consuming drugs’ and so no action was taken on not receiving a call. |
| Digital Pillbox | | |
| Trajman 2010 [23] | Obvious repetitive opening of the pillbox without pill withdrawal. | |
| Drabarek 2019 [24] | Digital data reports frequently underestimated adherence and masked the complications of separating the times at which the box was opened, and medication was taken. | |
| Video supported therapy | | |
| Hoffman 2010 [2] | | 50% of all anticipated videos were not received. |
| Lam 2018 [25] | | Out of 205 technical challenges experienced, 104/205 (51%) were related to patient adherence, including instances where patients were late or self-administered without provider approval. |
| Wade 2012 [9] | | The potential to cheat over the videophone was noticed and protocols instituted to minimize this. |

*Table 16: Decreased interactions affecting healthcare provider-patient relationship or treatment monitoring*

| Author Year | Findings |
| --- | --- |
| Video supported therapy | |
| Ritter 2017 [16] | Some reported that monitoring adverse effects of medications was more challenging using video supported therapy. |
| App-based | |
| Kopanitsa 2017 [20] | Doctors also mentioned that they were still responsible for the patients and would not like the portal to replace all the patients’ visits. |

#### Ease of use—Negative findings

*Table 17: Challenges contacting people with TB*

| Author Year | Findings |
| --- | --- |
| Phone-based | |
| Daftary 2017 [26] | No answer from patients (1056/2193 [48%]). Busy signal or the patient is on another call (76/2193 [4%]). |

#### Social influences—Negative findings

*Table 18: Decreased communication among healthcare providers*

| Author Year | Findings |
| --- | --- |
| 99DOTS | |
| Thekkur 2019 [22] | Lack of coordination between National TB Program and antiretroviral therapy clinic [i.e., HIV program] staff. |
| Prabhu 2021 [27] | Coordination challenges between TB and HIV center staff. |

#### Facilitating conditions—Negative findings

*Table 19: Suboptimal DAT function*

| Author Year | Findings |
| --- | --- |
| Short messaging service | |
| Hirsch-Moverman 2017 [5] | Health care providers also experienced technical challenges with the short messaging service; most were quickly resolved after discussions with study staff. |
| 99DOTS | |
| Thekkur 2019 [22] | Inability to add more than one mobile number per patient made engagement more challenging. |
| Thekkur 2019 [22] | Deficiencies in 99DOTS application: inability to register >1 phone per patient. |
| Thekkur 2019 [22] | Alert messages contained feedback in aggregate numbers rather than on details on individual patients who missed dose. |
| Thekkur 2019 [22] | Deficiency of adding treatment support on portal. |
| Prabhu 2021 [27] | Inability to add Revised National TB Control Programme staff as a treatment supporter in the system. |
| Prabhu 2021 [27] | Inadequate short messaging service alerts about non-adherent patients. |
| Prabhu 2021 [27] | Patient's missed calls did not get registered in the system. |
| Prabhu 2021 [27] | Software bugs limited use. |
| Thomas 2020 [3] | Patient's treatment status doesn't get updated in the dashboard. |
| Digital pillbox | |
| Thomas 2021 [14] | HCPs found intermittent (every 72 hours) updating of patients’ adherence records to be the most significant limitation to the Medication Event Reminder Monitor’s (MERM’s) perceived usefulness, as described by the following HCP: “*It takes 72 hours for the [MERM] dashboard to show that the patient has taken medications. This makes it difficult for us to monitor the patient’s drug intake on a daily basis. We cannot take action as promptly and lose time.*” [Pharmacist] |
| Video supported therapy | |
| Ritter 2017 [16] | Lost videos make it difficult to know whether patients have taken their medications or not. |
| Lam 2018 [25] | Out of 205 challenges experienced, 29/205 (14%) relate to health department equipment errors. |

*Table 20: Loss or destruction of the DAT*

| Author Year | Findings |
| --- | --- |
| Short messaging service | |
| Iribarren 2013 [28] | Temporary cellphone function problems (e.g., phone got wet or broke) affected reporting for some patients. |
| Digital Pillbox | |
| Trajman 2010 [23] | Loss or destruction of bottles was a problem. |
| Moulding 2002 [29] | 20% of medication monitors were not returned, of which 13% were lost by patients. |
| VOT | |
| Hoffman 2010 [2] | Of 50% of all anticipated videos that were not received, about 20% of those un-received videos were due to lost phones (1 stolen and 1 patient lost to follow-up). |

*Table 21: Inadequate Availability of Technology*

| Author Year | Finding |
| --- | --- |
| Digital pillbox | |
| Getachew 2022 [15] | Not enough computers. |

*Table 22: Complexity of cellular accessibility challenges (at a systemic level)*

| Author Year | Findings |
| --- | --- |
| Phone-based | |
| Bardosh 2017 [21] | Initial version in Kenya relied on local servers at clinics which were expensive and failed during power shortages. |
| 99DOTS | |
| Prabhu 2021 [27] | Network problems. |
| Thekkur 2019 [22] | The healthcare worker (HCW) had to log onto a portal which was difficult in the field (poor network coverage). |
| Digital pillbox | |
| Getachew 2022 [15] | Weak wifi signal. |
| Video supported therapy | |
| Wade 2012 [9] | Substantial and ongoing problems with video call quality were very frustrating. The call center nurses learned to manage most of these themselves. |
| Ritter 2017 [16] | Equipment and connectivity issues were challenges. |
| Ritter 2017 [16] | Numerous videos are received at once due to connectivity problems. |

*Table 23: Insufficient training or technology skills for healthcare providers*

| Author Year | | Findings |
| --- | --- | --- |
| Phone-based | | |
| Bardosh 2017 [21] | Low technology skills. | |
| 99DOTS | | |
| Thomas 2020 [3] | | Inadequate training of healthcare providers for health system changes |
| Thekkur 2019 [22] | | National TB Programme (NTP) staff not clear of their role and also who can be a treatment supporter. |
| Thekkur 2019 [22] | | Lack of training on new care package. |
| Thekkur 2019 [22] | | Lack of clarity on documentation of retrieval action after missed dose. |
| Prabhu 2021 [27] | | No formal training of staff in 99DOTS implementation. |
| Digital pillbox | | |
| Getachew 2022 [15] | | 26.7% of them were satisfied with the training given to them. Majority felt that the training provided was not enough to use the technology accordingly. |

*Table 24: Insufficient staff for DAT implementation*

| Author Year | Findings |
| --- | --- |
| Phone-based | |
| Bardosh 2017 [21] | Challenges for implementation and maintenance included: human resource shortages (initial study had dedicated staff paid at a higher rate that regular clinic staff, staff who were point persons were overburdened). |
| 99DOTS | |
| Thomas 2020 [3] | Increase in workload with 99DOTS implementation without changes in healthcare provider staffing. |
| Thekkur 2019 [22] | Antiretroviral Therapy clinic staff felt their workload had increased and National TB Programme (NTP) staff involvement was minimal (corroborated by TB staff). |
| Thekkur 2019 [22] | Inadequate human resources. |

*Table 25: Poor management of the DAT program*

| Author Year | Findings |
| --- | --- |
| 99DOTS | |
| Thekkur 2019 [22] | Lack of portal access to senior treatment supporter (STS). |
| Prabhu 2021 [27] | Coordination challenges between TB and HIV center staff. |
| Prabhu 2021 [27] | Inadequate supply of 99DOTS envelopes to facilitate implementation. |
| Thomas 2020 [3] | Technical problems with implementation (shortage of envelopes). |

*Table 26: Suboptimal culture of care*

| Author Year | Findings |
| --- | --- |
| Phone-based | |
| Bardosh 2017 [21] | Interactions between patients and providers were less frequent in Kenya due to less patient-centered care approaches. |
| Bardosh 2017 [21] | Low morale, work culture norm, low motivation for sustainment/maintenance once external support withdrawn; and lack of a local champion after WelTel staff were gone. |
| 99DOTS | |
| Thekkur 2019 [22] | Lack of ownership due to centralized care model; lack of role clarity; lack of involvement of general health system. |

*Table 27: Government barriers to implementation or sustainment*

| Author Year | Findings |
| --- | --- |
| Phone-based | |
| Bardosh 2017 [21] | Governments interested in using technology to solve existing care delivery problems rather than enhancing patient-centered care. |
| Bardosh 2017 [21] | Ongoing technical modification and redesign needed for ongoing maintenance (e.g., initial version in Kenya relied on local servers at clinics which were expensive and failed during power shortages, then switched to central server). |
| Bardosh 2017 [21] | WelTel team could not scale up without government co-financing, and governments would not finance without cost data -this dynamic was a barrier. |
| Video supported therapy | |
| Ritter 2017 [16] | Main barrier was that private insurance and public insurance do not reimburse for VDOT (public insurance does reimburse in-person DOT). |
| App-based | |
| Kopanitsa 2017 [20] | Some concern about the legal validity of the recommendations given through the portal. |

*Table 28: Equity challenges*

| Author Year | Findings |
| --- | --- |
| Video support therapy | |
| Ritter 2017 [16] | Connectivity problems are worse in rural regions. |

**References**

1.	Olano-Soler H, Thomas D, Joglar O, Rios K, Torres-Rodriguez M, Duran-Guzman G, et al. Notes from the Field: Use of Asynchronous Video Directly Observed Therapy for Treatment of Tuberculosis and Latent Tuberculosis Infection in a Long-Term-Care Facility - Puerto Rico, 2016-2017. MMWR Morb Mortal Wkly Rep. 2017;66: 1386–1387. doi:10.15585/mmwr.mm6650a5

2.	Hoffman JA, Cunningham JR, Suleh AJ, Sundsmo A, Dekker D, Vago F, et al. Mobile Direct Observation Treatment for Tuberculosis Patients. A Technical Feasibility Pilot Using Mobile Phones in Nairobi, Kenya. Am J Prev Med. 2010;39: 78–80.

3.	Thomas BE, Kumar JV, Onongaya C, Bhatt SN, Galivanche A, Periyasamy M, et al. Explaining Differences in the Acceptability of 99DOTS, a Cell Phone-Based Strategy for Monitoring Adherence to Tuberculosis Medications: Qualitative Study of Patients and Health Care Providers. JMIR MHealth UHealth. 2020;8: e16634.

4.	Mahmud N, Rodriguez J, Nesbit J. A text message-based intervention to bridge the healthcare communication gap in the rural developing world. Technol Health Care Off J Eur Soc Eng Med. 2010;18: 137–144. doi:10.3233/THC-2010-0576

5.	Hirsch-Moverman Y, Daftary A, Yuengling KA, Saito S, Ntoane M, Frederix K, et al. Using mhealth for HIV/TB treatment support in lesotho: Enhancing patient-provider communication in the start study. J Acquir Immune Defic Syndr. 2017;74: S37–S43.

6.	Thomas BE, Kumar JV, Onongaya C, Bhatt SN, Galivanche A, Periyasamy M, et al. Explaining Differences in the Acceptability of 99DOTS, a Cell Phone-Based Strategy for Monitoring Adherence to Tuberculosis Medications: Qualitative Study of Patients and Health Care Providers. JMIR MHealth UHealth. 2020;8: e16634.

7.	Wang N, Zhang H, Zhou Y, Jiang H, Dai B, Sun M, et al. Using electronic medication monitoring to guide differential management of tuberculosis patients at the community level in China. BMC Infect Dis. 2019;19: 844. doi:10.1186/s12879-019-4521-2

8.	Holzman SB, Zenilman A, Shah M. Advancing patient-centered care in tuberculosis management: A mixed-methods appraisal of video directly observed therapy. Open Forum Infect Dis. 2018;5: ofy046. doi:10.1093/ofid/ofy046

9.	Wade VA, Karnon J, Eliott JA, Hiller JE. Home Videophones Improve Direct Observation in Tuberculosis Treatment: A Mixed Methods Evaluation. PLoS ONE. 2012;7: e50155. doi:10.1371/journal.pone.0050155

10.	Iribarren SJ, Sward KA, Beck SL, Pearce PF, Thurston D, Chirico C. Qualitative evaluation of a text messaging intervention to support patients with active tuberculosis: implementation considerations. JMIR MHealth UHealth. 2015;3: e21. doi:10.2196/mhealth.3971

11.	Milligan H, Iribarren SJ, Chirico C, Telle H, Schnall R. Insights from participant engagement with the tuberculosis treatment support tools intervention: Thematic analysis of interactive messages to guide refinement to better meet end user needs. Int J Med Inf. 2021;149: 104421. doi:10.1016/j.ijmedinf.2021.104421

12.	Horter S, Stringer B, Venis S, Du Cros P. “I can also serve as an inspiration”: A qualitative study of the TB&Me blogging experience and its role in MDRTB treatment. PLoS ONE. 2014;9: e108591.

13.	Bardosh KL, Murray M, Khaemba AM, Smillie K, Lester R. Operationalizing mHealth to improve patient care: A qualitative implementation science evaluation of the WelTel texting intervention in Canada and Kenya. Glob Health. 2017;13: 87.

14.	Thomas BE, Kumar JV, Periyasamy M, Khandewale AS, Hephzibah Mercy J, Raj EM, et al. Acceptability of the Medication Event Reminder Monitor for Promoting Adherence to Multidrug-Resistant Tuberculosis Therapy in Two Indian Cities: Qualitative Study of Patients and Health Care Providers. J Med Internet Res. 2021;23: e23294.

15.	Getachew E, Woldeamanuel Y, Manyazewal T. Digital health interventions in the clinical care and treatment of tuberculosis and hiv in central Ethiopia: An initial provider perceptions and acceptability study using the unified theory of acceptance and use of technology model. Int J Mycobacteriology. 2022;11: 1–9.

16.	Ritter LA, Mei Wa K, Nasseri, Laura M. California Public Health Departments Remotely Treat Tuberculosis: Outcomes & Opportunities. Californian J Health Promot. 2017;15: 37–45.

17.	Story A, Aldridge RW, Smith CM, Garber E, Hall J, Ferenando G, et al. Smartphone-enabled video-observed versus directly observed treatment for tuberculosis: a multicentre, analyst-blinded, randomised, controlled superiority trial. The Lancet. 2019;393: 1216–1224. doi:10.1016/S0140-6736(18)32993-3

18.	Bendiksen R, Ovesen T, Asfeldt AM, Halvorsen DS, Gravningen K. Use of video call in the treatment of tuberculosis disease in Northern Norway. Tidsskr Nor Laegeforen. 2020;140: 35–40. doi:10.4045/tidsskr.19.0322

19.	Krueger K, Ruby D, Cooley P, Montoya B, Exarchos A, Djojonegoro BM, et al. Videophone utilization as an alternative to directly observed therapy for tuberculosis. Int J Tuberc Lung Dis. 2010;14: 779–781. doi:10.5588/ijtld.10.0210

20.	Kopanitsa G. A Qualitative Study of the Barriers and Opportunities for Adoption of Web-Portals for Doctors and Patients in Russia. J Med Syst. 2017;41: 62. doi:10.1007/s10916-017-0713-8

21.	Bardosh KL, Murray M, Khaemba AM, Smillie K, Lester R. Operationalizing mHealth to improve patient care: A qualitative implementation science evaluation of the WelTel texting intervention in Canada and Kenya. Glob Health. 2017;13: 87. doi:10.1186/s12992-017-0311-z

22.	Thekkur P, Kumar AMV, Chinnakali P, Selvaraju S, Bairy R, Singh AR, et al. Outcomes and implementation challenges of using daily treatment regimens with an innovative adherence support tool among HIV-infected tuberculosis patients in Karnataka, India: a mixed-methods study. Glob Health Action. 2019;12: 1–11. doi:10.1080/16549716.2019.1568826

23.	Trajman A, Long R, Zylberberg D, Dion MJ, Al-Otaibi B, Menzies D. Factors associated with treatment adherence in a randomised trial of latent tuberculosis infection treatment. Int J Tuberc Lung Dis. 2010;14: 551‐559.

24.	Drabarek D, Anh NT, Nhung NV, Hoa NB, Fox GJ, Bernays S. Implementation of Medication Event Reminder Monitors among patients diagnosed with drug susceptible tuberculosis in rural Viet Nam: A qualitative study. PloS One. 2019;14: e0219891. doi:10.1371/journal.pone.0219891

25.	Lam CK, McGinnis Pilote K, Haque A, Burzynski J, Chuck C, Macaraig M. Using Video Technology to Increase Treatment Completion for Patients With Latent Tuberculosis Infection on 3-Month Isoniazid and Rifapentine: An Implementation Study. J Med Internet Res. 2018;20: e287. doi:10.2196/jmir.9825

26.	Daftary A, Hirsch-Moverman Y, Kassie G, Melaku Z, Gadisa T, Saito S, et al. A Qualitative Evaluation of the Acceptability of an Interactive Voice Response System to Enhance Adherence to Isoniazid Preventive Therapy Among People Living with HIV in Ethiopia. AIDS Behav. 2017;21: 3057–3067.

27.	Prabhu A, Agarwal U, Singla N, Sarin R, Tripathy JP, Sagili K, et al. “99DOTS” techno-supervision for tuberculosis treatment - a boon or a bane? Exploring challenges in its implementation at a tertiary centre in Delhi, India. Indian J Tuberc. 2020;67: 46–53. doi:10.1016/j.ijtb.2019.08.010

28.	Iribarren S, Beck S, Pearce PF, Chirico C, Etchevarria M, Cardinale D, et al. TextTB: A Mixed Method Pilot Study Evaluating Acceptance, Feasibility, and Exploring Initial Efficacy of a Text Messaging Intervention to Support TB Treatment Adherence. Tuberc Res Treat. 2013;2013: 349394. doi:10.1155/2013/349394

29.	Moulding TS, Caymittes M. Managing medication compliance of tuberculosis patients in Haiti with medication monitors. Int J Tuberc Lung Dis. 2002;6: 313‐319. doi:10.5588/ijtld.02.0377
