## Supplemental Table 2 for "Contextual factors influencing implementation of tuberculosis digital adherence technologies: a scoping review guided by the RE-AIM framework": Supplementary Appendix 1 - Findings informing reach meta-themes.docx

**Findings informing “reach” meta-themes**

**Supplement to:**

**Contextual factors impacting implementation of tuberculosis digital adherence technologies: a scoping review using the RE-AIM framework**

S. Bahukudumbi, C. Chilala, B. Patel, N. Foster, M.S. Mohamed, M. Zary, C. Kafie, K. Schwartzman, K. Fielding, R. Subbaraman

**Correspondence:**

Ramnath Subbaraman, MD, MSc, FACP, FIDSA

Tufts University School of Medicine

Department of Public Health and Community Medicine

136 Harrison Ave., MV237

Boston, MA 02130, USA

### Reach (Positive contextual factors)

#### Perceived usefulness—Positive findings

*Table 1: Enhanced access to information and knowledge*

| Author Year | Findings |
| --- | --- |
| Short messaging service | |
| Nhavoto 2017 [1] | Participants thought that the system improves awareness of his/her disease. |
| Nhavoto 2017 [1] | 93% strongly agreed or agree that there are benefits of the short messaging service system in terms of receiving education and motivational messages. |
| Iribarren 2013 [2] | Educational text messages were helpful. |
| Video supported therapy | |
| Hoffman 2010 [3] | Patients welcomed health messaging as a part of the DAT intervention. |
| App-based | |
| Milligan 2021 [4] | App used to for disclosing treatment progress such as follow-up appointments, lab tests, and physician information and request assistance in making appointments for loved ones. |
| Iribarren 2020 [5] | App provided access to reliable TB information. |
| Iribarren 2020 [5] | App provided insights into one's medication dosing history. |
| Iribarren 2020 [5] | Allowed ability to record and view changes in side effects over time. |
| Iribarren 2020 [5] | App provided insights into one's medication dosing history. |

*Table 2: Improved medication adherence behavior*

| Author Year | Findings |
| --- | --- |
| Short messaging service | |
| de Sumari-de Boer 2016 [6] | Short messaging service reminder function was appreciated. |
| Nhavoto 2017 [1] | Participants thought that the short messaging service system helps patients to stay in treatment and increases treatment compliance. |
| Nhavoto 2017 [1] | 99% of participants strongly agreed or agreed that the system improves adherence to medication, reminding patients to collect medication on time. |
| Nhavoto 2017 [1] | 99% of participants strongly agreed or agreed that the short messaging service system improves adherence to medical appointments. |
| Khachadourian 2020 [7] | Text messages were perceived to be helpful in taking drugs every day for 78% participants. |
| Khachadourian 2020 [7] | Text messages were perceived to be helpful in taking drugs on time for 66% of participants. |
| Khachadourian 2020 [7] | Text message reminders were perceived to be helpful for visiting clinic weekly for 58% of participants. |
| Bediang 2018 [8] | Satisfaction Score: Support provided for adherence to drug prescriptions (99.6% of participants reported satisfaction on this item). |
| Iribarren 2013 [2] | Participants felt that short messaging service facilitated better adherence - that is, not forgetting to take medications because they know someone will be checking on them and reporting daily was "helpful". |
| Hirsch-Moverman 2017 [9] | 41.9% of patients stated the short messaging service messages were a facilitator to adherence in at least 1 monthly follow-up interview. |
| Phone-based | |
| Das Gupta 2020 [10] | Perceived live phone calls viewed as useful reminder cues, especially given work obligations. |
| Dessie Gashu 2020 [11] | Helped prevent patients from forgetting pills. |
| Daftary 2017 [12] | Patients felt benefit from daily reminders and greater awareness of pill-taking (habit formation). |
| Kopanitsa 2017 [13] | Patients felt inspired to follow doctors' recommendations, by using modern technologies. |
| 99DOTS | |
| Cross 2019 [14] | Timing of short messaging service reminder recognized as a trigger to take medications. |
| Thomas 2020 [15] | Reminders perceived by some to help adherence. |
| Digital pillbox | |
| van den Boogaard 2011 [16] | 68% of patients felt the MEMS bottle helped remind them to take medications. |
| Ratchakit-Nedsuwan 2020 [17] | Patients reported the reminder function was beneficial. |
| Bionghi 2018 [18] | Patients felt cared for (“felt important to have people worried about how I take medication") and this increased motivation to adhere. |
| Bionghi 2018 [18] | This form of monitoring encouraged them to take their medications at the prescribed times (45%). |
| Thomas 2021 [19] | Reminders promote medication adherence. |
| Video supported therapy | |
| Chen 2020 [20] | 100% strongly appreciate or appreciate efficacy to ensure treatment adherence. |
| Guo 2020 [21] | 80% of patients felt video observed therapy reduced the number of missed medication doses. |
| Guo 2020 [22] | Video directly observe therapy helped 92.9% of participants to not to miss doses. |
| Garfein 2015 [23] | 64% of patients found text message reminders helpful. |
| Hoffman 2010 [3] | Patients valued the reminders. |
| Holzman 2018 [24] | Video directly observe therapy provides improved adherence over self-administration (100% of participants). |
| Sinkou 2017 [25] | The high levels of adherence achieved; and the reduced risk of infecting other individuals during travel and clinic attendance. |
| App-based | |
| Navin 2017 [26] | 85% of patients felt the App was supportive for reminders and correct medicine intake. |

*Table 3: Improved monitoring of medication adverse effects*

| Author Year | Findings |
| --- | --- |
| Phone-based | |
| Kalita 2021 [27] | 80% of TB meningitis patients still on anti-TB therapy underwent therapy modification prompted by the phone outreach, of whom only 12% had to visit the health center. |
| Video supported therapy | |
| Chen 2020 [20] | 93% strongly appreciate or appreciate efficacy of video observed therapy for monitoring adverse events. |
| Olano-Soler 2017 [28] | Medical director observed patients over asynchronous videos for signs of hepatoxicity. |
| App-based | |
| Iribarren 2020 [5] | Allowed ability to record and view changes in side effects over time. |
| Milligan 2021 [4] | Improves monitoring of medication adverse effects. |

*Table 4: Storage and organization of medications*

| Author Year | Findings |
| --- | --- |
| Digital pillbox | |
| de Sumari-de Boer 2016 [6] | Storage function of the pillbox was appreciated. |
| Bionghi 2018 [18] | Helped participants to organize their medications (15%), and to keep their medications safe (45%). |
| Thomas 2021 [19] | Ease of pill taking because of better medication storage, organization, and labeling: “There are different compartments for each tablet, so they don’t get mixed with each other....It is helpful. I like the arrows with the dots that explain how many of each medication I need to take.” (Representative quotation from a 38-year-old male patient) |

*Table 5: Facilitates communication with providers*

| Author Year | Findings |
| --- | --- |
| Short messaging service | |
| Nhavoto 2017 [1] | Participants thought that the system improves communication between provider and patient. |
| Nhavoto 2017 [1] | 83% of participant reported great benefits or some perceived benefit of the short messaging service system for answering questions they sent. |
| Iribarren 2015 [29] | Providers felt TextTB was particularly helpful for communicating with, and supporting, TB patients in rural areas who were taking self-administered therapy. |
| Mahmud 2010 [30] | Remote requests to refill patient medications sent by providers (31% of messages facilitated patient referrals or reporting on patient issues). |
| Mahmud 2010 [30] | Notification of missed appointments by the patient are sent to the provider who can notify the patient (15% reported on patient symptoms). |
| Mahmud 2010 [30] | Patient queries can be reported via the healthcare worker to the doctor for rapid response. |
| Mahmud 2010 [30] | Requests for acute care can be communicated. |
| Iribarren 2013 [2] | Participants valued having someone available to answer their questions and the ability to consult with questions on side effects, TB transmission, etc. |
| Hirsch-Moverman 2017 [9] | Phone calls and above and beyond short messaging service messages empowered them to communicate with their health care providers and treatment supporters in a timely manner, without incurring a personal cost. Patients said they more frequently called their provider to report a side effect, seek advice, or inform their clinic about potential delays to a clinic appointment. They also felt more inclined to call and request assistance from their treatment supporters. |
| Phone-based | |
| Daftary 2017 [12] | Some participants found it easier and preferable to communicate with healthcare providers by phone and interactive voice response was appreciated to the extent it made it easier to connect with a healthcare provider. |
| Bardosh 2017 [31] | Facilitated greater access to care because barriers and wait times to reaching providers were reduced. |
| Bardosh 2017 [31] | Patients were assisted with emergencies through short messaging service texting. |
| Bardosh 2017 [31] | WelTel was perceived to empower patients to connect to medical staff. |
| Bardosh 2017 [31] | Facilitated greater access to care because barriers to reaching providers was lowered and wait times to reach providers reduced. |
| Bardosh 2017 [31] | Patients able to access assistance with emergencies through short messaging service texting. |
| Digital pillbox | |
| Ratchakit-Nedsuwan 2020 [17] | Patients report bi-directional communication with healthcare providers was beneficial. |
| Video supported therapy | |
| Olano-Soler 2017 [28] | Daily symptom queries attached to the videos may have informed care. |
| App-based | |
| Milligan 2021 [4] | App used to ask questions regarding TB treatment or personal circumstances such as medication phases, lab, and blood tests. |
| Iribarren 2020 [5] | App facilitated access to provide support to ask questions. |
| Navin 2017 [26] | 100% felt that the single touch for call the directly observed therapy provider was useful. |

*Table 6: Improved relationship between patient and provider*

| Author Year | Findings |
| --- | --- |
| 99DOTS | |
| Thomas 2020 [15] | Perceived improvements in the patient-healthcare provider relationship |
| Digital pillbox | |
| Thomas 2021 [19] | Strengthens patient-healthcare provider relationship. |
| Video supported therapy | |
| Wade 2012 [32] | Rapport with the nurses developed via video contact. |
| App Based | |
| Milligan 2021 [4] | Better relationships with patients. The treatment supporter emphasized the importance of building a trusting relationship with the participants and stated feeling “more like a treatment partner than a health professional conducting adherence surveillance.” |

*Table 7: Feeling of being cared for*

| Author Year | Findings |
| --- | --- |
| Short messaging service | |
| Gashu 2021 [33] | Reminders were useful and "supported me a lot." |
| Iribarren 2013 [2] | Short messaging service made participants feel “accompanied,” “cared for,” that they “had a friend when all others wanted nothing to do with them.” |
| Mohammed 2012 [34] | Five participants said they liked the messages because they found them encouraging and felt cared for. |
| Hirsch-Moverman 2017 [9] | Nurses and facility based voluntary health workers reported patients expressed feeling more cared for when they were followed up more frequently. Often, patients preferred to talk by phone rather than type a text message. |
| Phone-based | |
| Bardosh 2017 [31] | Facilitated a "sense that someone cares" for patients. |
| Daftary 2017 [12] | Patients felt grateful to be provided with mobile phones as otherwise they would not have had access to phones. |
| Daftary 2017 [12] | Patients felt "seen" by providers, especially because some had not disclosed their HIV to their family members. |
| 99DOTS | |
| Cross 2019 [14] | Monitoring fostered a sense of “responsibility and connection” with hospital staff. |
| Thomas 2020 [15] | Perceptions providers are using adherence data in a positive manner. |
| Digital pillbox | |
| Ratchakit-Nedsuwan 2020 [17] | Feeling of close proximity to the "doctor" (health professional). |
| Bionghi 2018 [18] | Patients felt cared for (“felt important to have people worried about how I take medication") and this increased motivation to adhere. |
| Thomas 2021 [19] | “If I don’t open the [pill] box, somebody from the health center calls me to find out whether I have taken the tablets or not. They care for me.” (Representative quotation from a male patient) |

*Table 8: Convenience of care delivery for people with TB*

| Author Year | Findings |
| --- | --- |
| Short messaging service | |
| Hermans 2017 [35] | Appointment reminders were generally found to be helpful; participants rated all the specific types of messages (reminder messages 94%, interactive quizzes 92%, and appointment reminders 94%) as equally helpful. |
| Iribarren 2015 [29] | TextTB could provide extra support during medication shortages, by letting patients know when medications had arrived. |
| Iribarren 2015 [29] | Providers felt TextTB was particularly helpful for communicating with, and supporting, TB patients in rural areas who were taking self-administered therapy. |
| Phone-based | |
| Bardosh 2017 [31] | Patients able to access assistance with emergencies through short messaging service texting. |
| Video supported therapy | |
| Wade 2012 [32] | More convenient scheduling was regarded as improving patient adherence. |
| Holzman 2018 [24] | 89% agreed with "video observed therapy is convenient". |

*Table 9: Convenience compared to alternative care models*

| Author Year | Findings |
| --- | --- |
| Short messaging service | |
| Iribarren 2013 [2] | Some patients preferred sending short messaging service over going to the clinic for dose observation. |
| Phone-based | |
| Bardosh 2017 [31] | Facilitated greater access to care because barriers to reaching providers was lowered and wait times to reach providers reduced. |
| Daftary 2017 [12] | Some participants found it easier and preferable to communicate with healthcare providers by phone and interactive voice response was appreciated to the extent it made it easier to connect with a healthcare provider. |
| 99DOTS | |
| Thomas 2020 [15] | Daily life not interrupted by facility-based directly observed therapy. |
| Digital pillbox | |
| Ratchakit-Nedsuwan 2020 [17] | Digital pillbox more convenient than in-person directly observed therapy at home, especially for evening dosing. |
| Thomas 2021 [19] | Reduces clinic visits. |
| Thomas 2021 [19] | Better medication storage, organization, and labeling: “Previously, I kept the tablets in a plastic cover, but now they are safer in the box. I used to be so confused, as there were so many medicines to take. Now it is easier.”; “The pills were previously given in an ordinary cardboard tablet box, which does not have an alarm, but this box has an alarm to remind me.” |
| Browne 2019 [36] | 100% of patients said they would prefer wireless observed therapy to directly observed therapy at randomization for the stage 2 trial. |
| Video supported therapy | |
| Bendiksen 2020 [37] | More convenient (“It was an advantage to have directly observed therapy by video conference. I felt I had more freedom, because I could take the drugs anywhere, and I could travel”). 82.4% preferred video conferencing. |
| Chen 2020 [20] | 89% strongly appreciate or appreciate convenience of flexibility in time scheduling. |
| Chen 2020 [20] | 100% strongly appreciate or appreciate convenience in location arrangement. |
| DeMaio 2001 [38] | Average "convenience" score of 8.8/10 points for use of video observed therapy in comparison to standard directly observed therapy. |
| Garfein 2015 [23] | 92% of patients prefer video directly observed therapy to in-person directly observed therapy. |
| Gassanov 2013 [39] | More efficient visits (reported by patients and staff). |
| Guo 2020 [21] | Participants undergoing video observed therapy felt that it was easier than going to the health center, was more likely to reduce missed doses, felt more satisfied with the model than with directly observed therapy. |
| Guo 2020 [22] | If a further treatment needed, 95.9% would choose the video directly observed therapy over another way. |
| Hoffman 2010 [3] | Percentage of patients who preferred video observed therapy over clinic-based or home-based directly observed therapy increased from 58% at start of study to 72% at the end of therapy. |
| Hoffman 2010 [3] | Agreement scores for preference for video observed therapy over directly observed therapy were relatively high at 4.6/5 at start of therapy and decline slightly to 4.2/5 at one month into therapy. |
| Holzman 2018 [24] | Video directly observe therapy increases access to transient patients. |
| Holzman 2018 [24] | Video directly observe therapy increases access to those with complicated work schedules. |
| Holzman 2018 [24] | Video directly observe therapy is more convenient for patients than directly observed therapy (88%). |
| Sinkou 2017 [25] | The improved convenience to patients of video observed therapy when compared with in-person direct observation. 80% of patients felt that video observed therapy was easier than daily commuting. |
| Sinkou 2017 [25] | Most welcomed the time and cost-saving attributes of video observed therapy. |
| Story 2019 [40] | Patients on directly observed therapy spent a mean 29 min (SD 48) per week on treatment observation (including travelling to or from clinics, waiting for appointments, and appointment time). Those on video observed therapy spent a mean of 1·8 min (2·2) setting up and recording each video. |
| Ting 2020 [41] | VDOT patients preferred video DOT over clinical-based DOT, having found it convenient and very accessible. The video phone application was easy to learn and connection was generally good. “Coming every day to the hospital is definitely more difficult than learning to connect to a conference on whatever a laptop, tablet. I mean, it’s pretty flexible to you, you just need an electronic device”. |
| Wade 2012 [32] | Patients could be observed at a time of their choosing, including early morning or evenings, fitting with lifestyle and cultural needs. |
| Wade 2009 [42] | One of the directly observed therapy clients was often hard to locate. “There is one client that is very challenging. It is very suitable for her because she is moving around so much, and it saves the round nurse chasing her down”. |
| Wade 2009 [42] | One of the two directly observed therapy clients was able to be called before work at 6.30AM and appreciated this flexibility of timing. |

*Table 10: Improving quality of care for people with TB*

| Author Year | Findings |
| --- | --- |
| Short messaging service | |
| Iribarren 2015 [29] | TextTB could provide extra support during medication shortages, by letting patients know when medications had arrived. |
| Phone-based | |
| Bardosh 2017 [31] | Patients could report malpractice or clinical errors, thereby improving care. |
| 99DOTS | |
| Thomas 2020 [15] | Perceived improvements in the quality of care. |
| Video supported therapy | |
| Holzman 2018 [24] | Video directly observe therapy was able to shorten therapy. |
| Hoffman 2010 [3] | Provides "a higher level of care" to patients. |
| Hoffman 2010 [3] | Allows for proactive and real time intervention on adverse effects and provision of counseling. |

#### Ease of use—Positive findings

*Table 11: DAT is easy to use*

| Author Year | Findings |
| --- | --- |
| Short messaging service | |
| Nhavoto 2017 [1] | 91% of participants strongly agreed or agreed that they had confidence in using the short messaging service system. |
| Nhavoto 2017 [1] | 77% of patients thought that the system was very easy or easy to use. |
| Digital pillbox | |
| Bionghi 2018 [18] | Acceptability toward the Wisepill device was strongly facilitated by its ease of use. |
| Bionghi 2018 [18] | The majority of participants encountered no difficulties removing the medications from the device every day. |
| Bionghi 2018 [18] | Most had no issue with this monitoring (35%). |
| Bionghi 2018 [18] | 95% found the technology easy to use; 5% found it not easy to use. |
| Ingestion sensors | |
| Belknap 2013 [43] | 83% of patients would be comfortable using ingestion sensors every day, and 75% would be comfortable using ingestion sensors in the long term. |
| Video supported therapy | |
| Holzman 2018 [24] | 100% found miDOT app easy to use, and 78% found video observed therapy easy to use. |
| DeMaio 2001 [38] | Average score of 8.4/10 points indicating video observed therapy was easy to master. |
| Guo 2020 [21] | 97% could record the video by themselves; 99% found video uploading to be very simple or simple; 935 found video uploading to be very easy or easy; 98% were satisfied with time spend on videos. |
| Guo 2020 [21] | 99% said the app caused problems during treatment <10% of the time; 99% were very satisfied or satisfied with the app interface. |
| Garfein 2020 [44] | 97% of video observed therapy users found it very easy or somewhat easy to use. |
| Garfein 2015 [23] | 92% of patients never/rarely had problems with recording videos. |
| Garfein 2015 [23] | 86% of patients never/rarely had problems with sending videos. |
| Ting 2020 [41] | Video observed therapy (video phone application) was easy to learn and connection was generally good. |
| Bommakanti 2020 [45] | 78.9% reported that the technology very easy to use; 17.9% reported that it was somewhat easy, and 3.3% reported that it was somewhat difficult. |
| Guo 2020 [22] | 95.9% of participants reported that the video directly observe therapy was convenient and comfortable. |
| App-based | |
| Iribarren 2020 [5] | Icons in app were easy to understand. |
| Navin 2017 [26] | 85% were able to use the App correctly. |
| Kopanitsa 2017 [13] | No comments on difficulties in using the portal. |

*Table 12: DAT provided flexibility and choice*

| Author Year | Findings |
| --- | --- |
| Video supported therapy | |
| Wade 2012 [32] | Chosen call time delivered reliably; patients did not need to wait. Patients could initiate a video observation when they were ready and change the time of observation at the last minute. Patients could move the videophone to another location whenever they chose. The technology was regarded as very easy to use. |
| Burzynski 2022 [46] | Flexibility in video observed therapy approach, with 42% choosing live-video electronic directly observed therapy; 42% choosing recorded-video electronic directly observed therapy; and the remainder choosing other directly observed therapy or self-administered options. |
| Olano-Soler 2017 [28] | Asynchronous video observed therapy facilitated observation successfully despite a poor internet connection, because it did not rely on a consistent connection and did not rely on a cellular plan. |

#### Social influences—Positive findings

*Table 13: Empowers or enhances autonomy for people with TB*

| Author Year | Findings |
| --- | --- |
| Short messaging service | |
| Iribarren 2013 [2] | Short messaging service made participants "feel responsible for their treatment." |
| Khachadourian 2020 [7] | Family supporters (80.8%) felt that the text messages helped them feel confident that the disease was under control. |
| Video supported therapy | |
| Holzman 2018 [24] | 79% agreed with the statement: "Video directly observe therapy provides autonomy." |
| App-based | |
| Horter 2014 [47] | The blogging experience was seen as giving patients voice (an empowerment tool). |
| Kopanitsa 2017 [13] | Patients reported that the portal made them feel involved in the treatment process and feel more responsibility for their health status. |

*Table 14: Low or reduced stigma from DAT use*

| Author Year | Findings |
| --- | --- |
| Short messaging service | |
| Hermans 2017 [35] | No participant reported adverse consequences as a result of other phone users reading their messages. None of the participants found the messages intrusive. |
| Hirsch-Moverman 2017 [9] | VHWs were sensitive to patients’ experiences with HIV-related stigma and supported the use of simple, coded text messages that protected patients’ privacy and confidentiality. |
| Digital pillbox | |
| Ratchakit-Nedsuwan 2020 [17] | Digital pillbox less of an invasion of privacy than in-person directly observed therapy at the home. |
| Bionghi 2018 [18] | No participant felt the device would invite stigma or a loss of confidentiality about their HIV or TB status. Indeed, several patients believed the device helped to maintain their confidentiality (30%). |
| Thomas 2021 [19] | Less stigma for patients (as reported by healthcare providers) compared to the prior box used for drug-resistant TB medications, because of no identifying writing on the outside of the DAT box. |
| Video supported therapy | |
| Holzman 2018 [24] | Video directly observe therapy is more private than standard directly observed therapy. |
| Chen 2020 [20] | 99% not at all concerned or unconcerned about privacy infringement with video observed therapy, as compared to 90% who were not at all concerned or unconcerned for standard directly observed therapy. |
| DeMaio 2001 [38] | Average score of 8.4/10 points indicated lower intrusiveness of video observed therapy in comparison to standard directly observed therapy. |
| Guo 2020 [21] | 57% of patients felt that uploading a video was never a privacy violation. |
| Garfein 2020 [44] | 62% of patients found video observed therapy to be more confidential that directly observed therapy. |
| Garfein 2015 [23] | 84% of patients felt video observed therapy was more confidential than in-person directly observed therapy and only 2% of patients failed to record a video because others may see them. |
| Wade 2012 [32] | Patients thought the videophone service was more private than an in-person service. |
| Bendiksen 2020 [37] | Confidentiality could best be upheld by video conferencing. |
| Gassanov 2013 [39] | Greater privacy (reported by patients and staff). |

*Table 15: Positive family involvement in care or social support*

| Author Year | Findings |
| --- | --- |
| Short messaging service | |
| Khachadourian 2020 [7] | Family supporters (80.8%) felt that the text messages helped them feel confident that the disease was under control. |
| Phone-based | |
| Kalita 2021 [27] | Caregivers often answered the phone with this intervention, so inclusion of caregivers may have facilitated ease of use and reach. |
| Bassett 2016 [48] | Individuals who were currently married (RR 2.1, 1.2-3.8) or who were never married (RR 1.8 ,1.0-3.3) had greater contact success. |
| 99DOTS | |
| Thomas 2020 [15] | Increased social support for some patients. |
| Digital pillbox | |
| Thomas 2021 [19] | Promotes family involvement in patient’s care. |
| Video supported therapy | |
| Garfein 2020 [44] | 74% of patients shared their video directly observe therapy experience with family members. |
| Garfein 2020 [44] | 34% shared their video directly observe therapy experience with friends, neighbors, schoolmates, workmates. |
| App-based | |
| Horter 2014 [47] | Blogging encouraged the patients to take their medications, partly not to disappoint those following the blog (social pressure). |
| Horter 2014 [47] | Blogging provided support to patients, with new peers resulting in reduction in isolation. |

#### Facilitating conditions—Positive findings

*Table 16: Adequate DAT training by providers enabled engagement*

| Author Year | Findings |
| --- | --- |
| Digital Pillbox | |
| Thomas 2021 [19] | Correct understanding of the Medication Event Reminder Monitor, reflecting appropriate counseling. |
| Video supported therapy | |
| Garfein 2015 [23] | 96% of patients found the video observed therapy training process helpful. |

*Table 17: Provision of phone enabled DAT engagement*

| Author Year | Findings |
| --- | --- |
| Video supported therapy | |
| Garfein 2020 [44] | 39% of patients had to have a smartphone loaned to them to facilitate video observed therapy and this was done. |

*Table 18: Optimal DAT Function or DAT Problems Easily Resolved*

| Author Year | Findings |
| --- | --- |
| Short messaging service | |
| Liu 2015 [49] | 89% of mobile phone problems were resolvable. |
| Video supported therapy | |
| Wade 2012 [32] | Occasional whole of system failures occurred but were deemed manageable. |
| Olano-Soler 2017 [28] | Asynchronous video observed therapy facilitated observation successfully despite a poor internet connection, because it did not rely on a consistent connection and did not rely on a cellular plan. |
| Garfein 2015 [23] | Improvement in comfort using smartphone to record videos by 1.68/10 points over treatment. |
| Garfein 2015 [23] | 92% of patients never/rarely had problems with recording videos. |
| Garfein 2015 [23] | 86% of patients never/rarely had problems with sending videos. |

*Table 19: No or only minor challenges with network function*

| Author Year | Findings |
| --- | --- |
| Video supported therapy | |
| Garfein 2015 [23] | 96% of patients never/rarely had problems with sending videos due to poor reception. |
| Wade 2012 [32] | Occasional whole of system failures occurred but were deemed manageable. |

### Reach (Negative contextual factors)

#### Perceived usefulness—Negative findings

*Table 20: Greater access to information desired*

| Author Year | Findings |
| --- | --- |
| Short messaging service | |
| Iribarren 2013 [2] | Some participants wanted even more information about TB to be provided. |
| App-based | |
| Iribarren 2020 [5] | Listing of health centers within app was desired. |
| Iribarren 2020 [5] | TB information could have been better delivered through videos rather than text. |

*Table 21: DAT does not address patient problems*

| Author Year | Findings |
| --- | --- |
| Phone-based | |
| Das Gupta 2020 [10] | Providers felt DAT did not address main causes of medication non-adherence, which were medication adverse effects and alcohol use. |
| Bardosh 2017 [31] | Short messaging service texting was of limited use for addressing social determinants (e.g., poverty) and structural barriers (e.g., drug stock outs) faced by patients, particularly in Kenya and tech intervention were not designed to connect them to social services. |
| Dessie Gashu 2020 [11] | Only 17% of patients were taking medications at suggested times, raising questions about utility of DAT for improving adherence. |

*Table 22: Technology fatigue*

| Author Year | Findings |
| --- | --- |
| Short messaging service | |
| Gashu 2021 [33] | Some patients feel "bored" with [SMS text] messages and they create unpleasant moods. |
| Iribarren 2013 [2] | Some patients felt lack of motivation to send messages daily and requested less frequent notification ("technology fatigue"). |
| 99DOTS | |
| Thomas 2020 [15] | Perceived high burden of calling. |
| Thekkur 2019 [50] | Lack of motivation to give a missed call [i.e., call 99DOTS]. |
| Thekkur 2019 [50] | Failing to give missed call, lack of motivation for giving missed call. |
| Prabhu 2021 [51] | Lack of motivation to call. |

*Table 23: DAT had physical adverse effects*

| Author Year | Finding |
| --- | --- |
| Ingestion Sensor | |
| Browne 2019 [36] | 9.8% of study participants had mild grade 1 reactions to the patch they had to wear, and one patient withdrew from the study for this reason. |

*Table 24: Different or modified DAT modality desired*

| Author Year | Findings |
| --- | --- |
| Short messaging service | |
| de Sumari-de Boer 2016 [6] | Patients desired that an additional short messaging service reminder be sent before the actual time of medication intake. |
| Gashu 2021 [33] | Many patients received the short messaging service reminder after taking pills. |
| Gashu 2021 [33] | Some patients wanted short messaging service to go beyond reminders to address nutrition, drug side effects, lab tests, TB knowledge, and motivational messages. |
| Iribarren 2013 [2] | Some people wanted to be able to email to report doses. |
| Video supported therapy | |
| Hoffman 2010 [3] | Patients preferred personal testimonial videos as the best method for communicating messages regarding TB as part of DAT intervention. |
| App-based | |
| Iribarren 2020 [5] | TB information could have been better delivered through videos rather than text. |
| Iribarren 2020 [5] | Communication via a known app such as WhatsApp thought to be more useful given prior knowledge of app by patients. |
| Iribarren 2020 [5] | Reminder function within app was desired. |

*Table 25: Purpose of DAT monitoring was unclear*

| Author Year | Findings |
| --- | --- |
| 99DOTS | |
| Thomas 2020 [15] | Lack of clarity on what 99DOTS is or its purpose in their care. |
| Thekkur 2019 [50] | Lack of awareness regarding need for giving missed call. |

*Table 26: Negative impact on relationship with healthcare system*

| Author Year | Findings |
| --- | --- |
| 99DOTS | |
| Thomas 2020 [15] | Technology negatively impacts patient-provider relationship. |
| Thomas 2020 [15] | Inadequate or negatively perceived outreach by providers. |
| Thekkur 2019 [50] | Perception that centralized TB treatment at ART center without involving general health system or TB program staff had reduced the number of the patient–provider interactions and leading to an increase in the “loss to follow-up”. |
| Digital pillbox | |
| Drabarek 2019 [52] | Despite research staff receiving adherence data from the devices, patients in the intervention group did not report that this information had been raised in their discussions with a health worker. |
| Video supported therapy | |
| Gassanov 2013 [39] | Diminished interpersonal interaction (reported by patients and staff). |

*Table 27: Preference for in-person healthcare provider-patient communication*

| Author Year | Findings |
| --- | --- |
| Short messaging service | |
| Iribarren 2013 [2] | Some participants wanted more in-person contact in addition to texting or to be called rather than texted for more personal contact. |
| Phone-based | |
| Daftary 2017 [12] | A smaller group of participants strongly preferred interacting with healthcare providers in person rather than over the phone. |
| App-based | |
| Kopanitsa 2017 [13] | Patients expected a 24/7 direct communication with the doctor after each data exchange. |

*Table 28: Concerns about surveillance*

| Author Year | Findings |
| --- | --- |
| Digital pillbox | |
| Bionghi 2018 [18] | Fear of missing a dose (5%). Some expressed fear of the monitoring ("If you miss the correct time, you become very scared.") |
| Thomas 2021 [19] | Perceptions that the Medication Event Reminder Monitor facilitated government surveillance: ““There is a camera in the box, so if you don’t take the pills, people in Delhi will come to know. So take your pills.” (Mother of a 25-year-old male patient) |

#### Ease of use—Negative findings

*Table 29: Complexity of cellular accessibility challenges (at an individual level)*

| Author Year | Findings |
| --- | --- |
| Short messaging service | |
| Mohammed 2012 [34] | 17.85% didn't respond due to cell-phone problems (running out of phone credit, the line being switched off, and the phone’s battery running out and losing the mobile phone). |
| Liu 2015 [49] | Lack of money for air time: among 42% of patients who experienced DAT problems in the short messaging service only trial arm, 15% of all problems were due to lack of money for air time. |
| Liu 2015 [49] | Incorrect phone used: among 42% of patients who experienced mobile phone problems, 42% of all problems involved using an incorrect phone and not receiving short messaging service texts. |
| Iribarren 2015 [29] | Adding phone credit was time consuming and credit was depleted early for some participants. |
| Iribarren 2013 [2] | Shared cellphone use was a barrier to engagement for 3/18 (17%) of patients with two of these patients having low engagement (<50% of doses reported via short messaging service). |
| Iribarren 2013 [2] | Running out of cellphone credit was a temporary barrier for some. |
| Mahmud 2010 [30] | Of all messages, 219/1330 (17%) were to request short messaging service credit, and 28/1330 (2%) were due to phone problems. |
| Hirsch-Moverman 2017 [9] | A few patient participants did not use or own a phone. They had registered their treatment supporter to receive short messaging service reminders, who in turn, reminded them to take medications. As a result, they did not fully understand or appreciate the utility of the short messaging service. |
| Phone-based | |
| Bardosh 2017 [31] | Only 50% enrolled in some locations in Kenya due to low literacy and lack of cellphone ownership. |
| Bardosh 2017 [31] | Access to short messaging service texting believed to be lower among Kenyan pastoralists and women in Kenya. |
| 99DOTS | |
| Thekkur 2019 [50] | Lack of family support and reliance on a family member's phone who was not available. |
| Thekkur 2019 [50] | Failing to give missed call. Patients don't own phone and have to rely on family members. |
| Prabhu 2021 [51] | Mobile phones are unaffordable, so access is poor. |
| Prabhu 2021 [51] | Shared cellphone use with family members limited ability to call. |
| Video supported therapy | |
| Buchman 2017 [53] | Dependency on an unreliable family member for Skype access. |
| Bommakanti 2020 [45] | Not owning a smartphone was associated with increasing age, male sex, high school education or below (compared to above high school), and income <$10,000 per year. |
| Gassanov 2013 [39] | Physical assessment over a videophone can be difficult, particularly if picture quality is poor; most technical issues were concentrated among a few patients (suggesting the quality of the patients’ internet connections is the issue). |
| Olano-Soler 2017 [28] | Lack of cellphones and poor internet connectivity made live video directly observed therapy infeasible in the long-term care facility. |
| Ting 2020 [41] | Half of patients reported that the fixed video appointment time was still hard to meet especially during work hours. The fixed video appointment requires real-time cellular connection and is only feasible if patients have access to a suitable device and stable internet connection. |

*Table 30: Literacy and language barriers*

| Author Year | Findings |
| --- | --- |
| Short messaging service | |
| Cox 2018 [54] | 34% of TB patients in two settings felt uncomfortable sending and receiving short messaging service (57% in India and 8% in South Africa). |
| Hermans 2017 [35] | A quarter of participants stated they did not always understand the messages due to the technical language used (28%). |
| Mohammed 2012 [34] | Two participants said that they would prefer it if the messages were in Urdu script, rather than Urdu transliterated into English script, as they were not familiar with the latter and were not always able to understand the messages. |
| Mohammed 2012 [34] | 2/28 didn’t respond because they couldn't read and there was no one at home who could read at the time. |
| Mohammed 2012 [34] | Most women relied on male family members to help with reading messages and responding. |
| Phone-based | |
| Daftary 2017 [12] | Some participants found it hard to understand and respond to automated messages. |
| Bardosh 2017 [31] | Only 50% enrolled in some locations in Kenya due to low literacy and lack of cellphone ownership. |
| 99DOTS | |
| Cross 2019 [14] | Lack of understanding of content of reminders. |
| Thomas 2020 [15] | Short messaging service texts are in a language patient does not understand. |
| Thomas 2020 [15] | Patient ease of use was perceived to be variable, depending on educational level. |
| Thekkur 2019 [50] | Lack of capacity [of patients] to operate mobile phone. |
| Thekkur 2019 [50] | Did not know how to use a phone. |
| App-based | |
| Iribarren 2020 [5] | Some app items remained in English making them difficult to read. |
| Kopanitsa 2017 [13] | Issues with understanding recommendations from the doctor. Patient needed to ask advice from friends/relatives to understand recommendations. |

*Table 31: DAT too technically complex or physically difficult to use*

| Author Year | Findings |
| --- | --- |
| Short messaging service | |
| Hermans 2017 [35] | Many commented on the complexity of the messages and their frustration at trying to send a reply that was not recognized by the system. |
| Hirsch-Moverman 2017 [9] | Patients who were generally less familiar with mobile phones were at times confused by the coded messages. Nonetheless, they managed to link the short messaging service messages to medication intake. |
| 99DOTS | |
| Thomas 2020 [15] | Envelope was too complex, leading to inappropriate calling. |
| Thekkur 2019 [50] | Cumbersome for some patients to regularly type a 10-digit number for a missed call. |
| Thekkur 2019 [50] | Problems with calling: difficulty for patients in dialing the 10-digit number and absence of automated voice reply (made it difficult to know if the call was registered). |
| Prabhu 2021 [51] | Many patients were unable to operate mobile phones. |
| Digital pillbox | |
| Ratchakit-Nedsuwan 2020 [17] | Smart phones were too difficult to operate for the elderly. |
| Ratchakit-Nedsuwan 2020 [17] | Instructions on the box were too small to read. |
| Bionghi 2018 [18] | A small number (15%) of participants with stiff joints experienced some difficulty opening the device and one (5%) participant required an extra demonstration to master handling the device. |
| Video supported therapy | |
| Chuck 2016 [55] | Patient out of camera view: 80% of patients experienced at least one video observed therapy issue, of which 27% involved the patient being out of camera view. |
| Chuck 2016 [55] | Challenges using smartphone: 80% of patients experienced at least one video observed therapy issue, of which 26% involved patient not being able to operate smartphone, broken accessories, or blocking of the camera. |
| App-based | |
| Milligan 2021 [4] | Missed report of urine metabolite test strip result due to reporting in a WhatsApp message rather than secure App, change in life routines due to medication side effects or work routine, forgetting to report, and unforeseen life events. |
| Milligan 2021 [4] | Many communications related to request for assistance with App issues, request to confirm receipt of urine test result, and assistance with the overall intervention. |
| Iribarren 2020 [5] | Difficulties with uploading of urine test photos. |

*Table 32: Size of DAT is prohibitive*

| Author Year | Findings |
| --- | --- |
| Digital pillbox | |
| van den Boogaard 2011 [16] | 22% of patients took medications out of the MEMS for later use, often due to travel. |
| de Sumari-de Boer 2016 [6] | When patients travelled pillbox was felt to be obtrusive (too large). |
| Drabarek 2019 [52] | Box was too big and lacked practical portability; participants preferred to take multiple pills out at a time rather than open the box daily. Pill were removed to facilitate pill-taking within the patient's existing routine (e.g., taking them at work). |
| Thomas 2021 [19] | As reported by healthcare providers: The size of the Medication Event Reminder Monitor made transporting the device to clinic visits prohibitive. |
| Thomas 2021 [19] | Size, portability, and storage problems. |

#### Social influences—Negative findings

*Table 33: High or increased stigma*

| Author Year | Findings |
| --- | --- |
| Short messaging service | |
| Iribarren 2013 [2] | Stigma influenced ability to engage for some patients ("yes many people are even scared to call"). |
| Mohammed 2012 [34] | A participant was concerned about being stigmatized if anyone came across the message. |
| Phone-based | |
| Daftary 2017 [12] | Receiving a phone became a marker of patients' illness and some were afraid that it would lead to disclosure of their condition (HIV). |
| Bardosh 2017 [31] | Receiving a phone became a marker of patients' illness and some were afraid that it would lead to disclosure of their condition (HIV). |
| 99DOTS | |
| Thomas 2020 [15] | DAT or provider response to engagement data increased anticipated or experienced stigma. |
| Thekkur 2019 [50] | Stigma may have contributed to avoidance of house visits by providers, which is a recommended follow-up action based on 99DOTS data. |
| Thekkur 2019 [50] | Lack of family support: stigma and avoidance of house visit by providers. |
| Digital pillbox | |
| Ratchakit-Nedsuwan 2020 [17] | Patients were stigmatized. |
| de Sumari-de Boer 2016 [6] | Had to hide box in case household members opened it, thereby causing issues with data (of box opening). |
| Thomas 2021 [19] | Problems related to privacy and stigma due to the size and visibility of the box and the loudness of the reminder alarm. |
| Thomas 2021 [19] | Audible reminder is too loud. |
| Video supported therapy | |
| Guo 2020 [21] | 43% of patients felt that uploading a video was occasionally, often, or always a privacy violation, at a similar level to perceptions of privacy violation with standard directly observed therapy. |
| Chuck 2016 [55] | Lack of privacy: 80% of patients experienced at least one video observed therapy issue, of which 37% were related to patients forgetting medications at home, forgetting appointments, or being unable to find private place. |
| Garfein 2020 [44] | 40% of patients concerned someone would see them doing video observed therapy and 10% did not record videos for this reason. |
| Wade 2012 [32] | Two patients expressed privacy concerns. |
| Holzman 2018 [24] | Video directly observe therapy can compromise patient privacy 2/16 (13%). |
| Nhavoto 2017 [1] | Only 9% of participants agreed or strongly agreed that there were privacy or confidentiality risks with the system. |
| Holzman 2019 [56] | Confidentiality concerns with video directly observe therapy 3/10 (30%) impacted participants’ ability to make it through the 10 day run-in period. |
| App-based | |
| Iribarren 2020 [5] | Use of "TB" in app name thought to be stigmatizing and put patients at risk of disclosure. |

*Table 34: Desire for greater social contact via DAT with other people with TB*

| **Author Year Citation** | **Findings** |
| --- | --- |
| App Based | |
| Iribarren 2020 [5] | Communication function with other TB patients was desired. |

*Table 35: Concerns about family involvement in care*

| Author Year | Findings |
| --- | --- |
| 99DOTS | |
| Thekkur 2019 [50] | Lack of family support and reliance on a family member's phone who was not available. |
| Digital pillbox | |
| Ratchakit-Nedsuwan 2020 [17] | Concern that small children will play with the pillbox. |
| Thomas 2021 [19] | Concern about children playing with the pillbox: ““*I keep my box in a hen cage [outside of the house], because my children used to play with it. I don’t have a place in my home to keep the box where my children won’t reach it*.” (Patient, male, aged 33 years) |
| Video supported therapy | |
| Buchman 2017 [53] | Dependency on an unreliable family member for Skype access. |

*Table 36: Social events or holidays reduce DAT engagement*

| Author Year | Findings |
| --- | --- |
| Digital Pillbox | |
| Stagg 2020 [57] | Lower engagement with the MERM was associated with weekends, national holidays and transition from the intensive to the continuation phase of therapy in an adjusted regression analysis. |

*Table 37: Socially desirable behavior to please provider leads to inaccurate adherence data*

| Author Year | Findings |
| --- | --- |
| Digital Pillbox | |
| Trajman 2010 [58] | Obvious repetitive opening of the pillbox without pill withdrawal. |

*Table 38: Concerns about high monetary value of the DAT*

| Author Year | Findings |
| --- | --- |
| Digital Pillbox | |
| de Sumari-de Boer 2016 [6] | Concerns digital pillbox might be stolen as it looks like a camera. |

#### Facilitating conditions—Negative findings

*Table 39: Suboptimal DAT Function*

| Author Year | Findings |
| --- | --- |
| Short messaging service | |
| Iribarren 2015 [29] | Laptop powering down sent messages out of sync with expected timing, confusing participants. |
| Phone Based | |
| Daftary 2017 [12] | PIN [i.e., personal identification number to hear interactive voice recordings] failure for 304/2193 (14%) patients. |
| 99DOTS | |
| Thekkur 2019 [50] | When calling 99DOTS, call did not seem to connect on the other end. |
| Thekkur 2019 [50] | Problems with calling: difficulty for patients in dialing the 10-digit number and absence of automated voice reply [made it difficult to know if the call was registered]. |
| Thekkur 2019 [50] | Patients expected that someone would talk to them upon giving call to the 99DOTS portal and this led to confusion. |
| Thomas 2020 [15] | Patient received busy signal when calling 99DOTS: “*The first time I call, it gives a busy signal . . . after thinking that I dialed a wrong number, I dial the number again and it works... this has happened two or three times*.” (43 year-old-woman, HIV coinfected, intensive phase of therapy) |
| Digital pillbox | |
| Trajman 2010 [58] | Problems with the electronic device. |
| Drabarek 2019 [52] | Alarm was redundant, as MERM alarm timing not aligned with when patients were told to take medications by the doctor, leading patients to take pills out to ingest at another time. |
| Ratchakit-Nedsuwan 2020 [17] | Patients complained about fast battery draining. |
| Ratchakit-Nedsuwan 2020 [17] | Inaudible alarm when placed on another floor in the house. |
| Thomas 2021 [19] | Failure of the device's battery. |
| Thomas 2021 [19] | Malfunction of reminder lights (all lights were glowing continuously). |
| Thomas 2021 [19] | Audible reminder is too loud. |
| Video supported therapy | |
| Wade 2012 [32] | Substantial and ongoing problems with video call quality were very frustrating. |
| Hoffman 2010 [3] | Videos occasionally required resending due to network congestion. |
| Hoffman 2010 [3] | Of 50% of all anticipated videos that were not received, about 50% of those videos that were not received were due to technical issues preventing transmission and 30% were due to unknown reasons. |
| Lam 2018 [59] | Out of 205 challenges experienced, 29/205 (14%) relate to health department equipment errors. |
| App-based | |
| Milligan 2021 [4] | App access difficulty due to phone problems or inconsistent Wi-Fi; App not functioning; App says urine test result was reported but it does not get recorded in App. |
| Milligan 2021 [4] | Missed urine metabolite test strip result report due to: App access difficulty due to phone problems or inconsistent Wi-Fi; App not functioning; App says urine test result was reported but it does not get recorded in App. |

*Table 40: Complexity of cellular accessibility challenges (at a systemic level)*

| Author Year | Findings |
| --- | --- |
| Short messaging service | |
| Gashu 2021 [33] | Short messaging service reminders were completely interrupted for a few days by telecom company due to large volume of messages from a single provider. |
| Gashu 2021 [33] | Some participants reported lack of electric power or poor telecommunication network connections as barriers. |
| Mohammed 2016 [60] | 21% of short messaging service reminders were not sent due to system failures, GPRS outages, and administrative shortfalls |
| Hermans 2017 [35] | Messages were not delivered to participants when a particular network provider had interrupted service. |
| Liu 2015 [49] | Network failure: Among 42% of patients who experienced DAT problems in the short messaging service only trial arm, 21% of all problems were due to network failure. |
| de Sumari-de Boer 2016 [6] | Short messaging service reminders sent in error due to network issues. |
| Phone-based | |
| Dessie Gashu 2020 [11] | Inconsistency of power supply. |
| Dessie Gashu 2020 [11] | Weak mobile network. |
| Daftary 2017 [12] | Failed interactive voice response call due to network malfunction (540/2193 [25%]). |
| Hirsch-Moverman 2017 [9] | Unstable access to electricity and temporary technical difficulties appeared to bar a few patient participants from receiving regular short messaging service reminders. |
| 99DOTS | |
| Thekkur 2019 [50] | Poor network coverage was a barrier to 99DOTS engagement. |
| Thomas 2020 [15] | Lack of electricity in the home. |
| Thomas 2020 [15] | Lack of cellular network in the home. |
| Digital Pillbox | |
| Ratchakit-Nedsuwan 2020 [17] | Half of patients had poor signal or lack of mobile signal. |
| Thomas 2021 [19] | Person with TB was being called by healthcare provider because doses were not being reported due to network problems. |
| Video supported therapy | |
| Olano-Soler 2017 [28] | Lack of cellphones and poor internet connectivity made live video directly observed therapy infeasible in the long-term care facility. |
| Ting 2020 [41] | Half of patients found that the fixed video appointment time was still hard to meet especially during work hours. The fixed video appointment time required real-time cellular connection and is only feasible if patients have access to a suitable device and stable internet connection. |
| Ratchakit-Nedsuwan 2020 [17] | Half of patients had poor signal or lack of mobile signal. |
| Holzman 2019 [56] | Unable to consistently submit videos for 1/10 participants (10%), inconsistent internet or wifi for 1/10 participants (10%). |

*Table 41: Lack of adequate counseling on DAT*

| Author Year | Findings |
| --- | --- |
| 99DOTS | |
| Thomas 2020 [15] | Poor counseling in purpose or use of DAT. |
| Thekkur 2019 [50] | Lack of awareness regarding need for giving missed call. |
| Thekkur 2019 [50] | Fear of being charged for missed call [represents incorrect knowledge of the intervention]. |
| Thekkur 2019 [50] | Misperception that a person would talk to them upon calling 99DOTS. |
| Prabhu 2021 [51] | Toll-free number concept was misunderstood [perhaps lack of counseling]. |
| Digital Pillbox | |
| Thomas 2021 [19] | Incorrect understanding of the Medication Event Reminder Monitor, reflecting suboptimal counseling. |

*Table 42: Suboptimal culture or quality of general TB care leads to poor DAT implementation*

| Author Year | Findings |
| --- | --- |
| Short messaging service | |
| Iribarren 2015 [29] | Medication shortages meant that medications could not be dispensed to patients in a manner concordant with the short messaging service intervention. |
| Phone-based | |
| Bardosh 2017 [31] | Interactions between patients and providers were less frequent in Kenya due to less patient-centered care approaches. |
| Digital pillbox | |
| Thomas 2021 [19] | MDR-TB medications were supposed to be dispensed in the MERM on a monthly basis; however, some medications were sometimes understocked. . . MERM implementation therefore worsened challenges related to the understocking of drugs: “*Sometimes MDR-TB drugs are not available, and so we are not able to give all the medicines required....How do we leave that compartment [in the MERM for a specific medication] empty, and what can we tell the patient?”* [Pharmacist] |

*Table 43: Equity challenges*

| Author Year | Findings |
| --- | --- |
| Short messaging service | |
| Liu 2015 [49] | Lack of money for airtime: among 42% of patients who experienced DAT problems in the short messaging service only trial arm, 15% of all problems were due to lack of money for airtime. |
| Mohammed 2012 [34] | Two participants said that they would prefer it if the messages were in Urdu script, rather than Urdu transliterated into English script, as they were not familiar with the latter and were not always able to understand the messages. |
| Mohammed 2012 [34] | Most women relied on male family members to help with reading messages and responding. |
| Bommakanti 2020 [45] | Not owning a smartphone was associated with increasing age, male sex, high school education or below (compared to above high school), and income <$10,000 per year. |
| Phone-based | |
| Person 2011 [61] | Younger individuals were more likely to have cellphone access and those with higher education were more likely to have cellphone access. |
| Bassett 2016 [48] | Individuals with 0 barriers to care (RR 1.6, 1.3-2.05) or 1-3 barriers to care (1.3, 1.0-1.7) were more likely to achieve >=5 calls (presumably compared to those with 4 or more barriers to care) [indicates that already disadvantaged patients may have greater challenges engaging with DATs]. |
| Bardosh 2017 [31] | Access to short messaging service texting believed to be lower among Kenyan pastoralists and women in Kenya. |
| Bardosh 2017 [31] | Only 50% enrolled in some locations in Kenya due to low literacy and lack of cellphone ownership. |
| 99DOTS | |
| Prabhu 2021 [51] | Mobile phones are unaffordable, so access is poor. |
| Thomas 2020 [15] | Inability to read the number on the 99DOTS envelope: “*My vision is not clear enough to see the small print* [on the envelope]*... I get help from my daughter or someone who can see the letters and call*.” (42-year-old man, HIV coinfected, continuation phase of therapy) |
| Digital pillbox | |
| Bionghi 2018 [18] | A small number (15%) of participants with stiff joints experienced some difficulty opening the device and one (5%) participant required an extra demonstration to master handling the device. |

**References**

1.	Nhavoto JA, Grönlund Å, Klein GO. Mobile health treatment support intervention for HIV and tuberculosis in Mozambique: perspectives of patients and healthcare workers. PloS ONE. 2017;12: e0176051. doi:10.1371/journal.pone.0176051

2.	Iribarren S, Beck S, Pearce PF, Chirico C, Etchevarria M, Cardinale D, et al. TextTB: A Mixed Method Pilot Study Evaluating Acceptance, Feasibility, and Exploring Initial Efficacy of a Text Messaging Intervention to Support TB Treatment Adherence. Tuberc Res Treat. 2013;2013: 349394. doi:10.1155/2013/349394

3.	Hoffman JA, Cunningham JR, Suleh AJ, Sundsmo A, Dekker D, Vago F, et al. Mobile Direct Observation Treatment for Tuberculosis Patients. A Technical Feasibility Pilot Using Mobile Phones in Nairobi, Kenya. Am J Prev Med. 2010;39: 78–80.

4.	Milligan H, Iribarren SJ, Chirico C, Telle H, Schnall R. Insights from participant engagement with the tuberculosis treatment support tools intervention: Thematic analysis of interactive messages to guide refinement to better meet end user needs. Int J Med Inf. 2021;149: 104421. doi:10.1016/j.ijmedinf.2021.104421

5.	Iribarren SJ, Rodriguez Y, Lin L, Chirico C, Discacciati V, Schnall R, et al. Converting and expanding a mobile support intervention: Focus group and field-testing findings from individuals in active tuberculosis treatment. Int J Med Inf. 2020;136: 104057.

6.	de Sumari-de Boer IM, van den Boogaard J, Ngowi KM, Semvua HH, Kiwango KW, Aarnoutse RE, et al. Feasibility of Real Time Medication Monitoring Among HIV Infected and TB Patients in a Resource-Limited Setting. AIDS Behav. 2016;20: 1097–1107. doi:10.1007/s10461-015-1254-0

7.	Khachadourian V, Truzyan N, Harutyunyan A, Petrosyan V, Davtyan H, Davtyan K, et al. People-centred care versus clinic-based DOT for continuation phase TB treatment in Armenia: a cluster randomized trial. BMC Pulm Med. 2020;20: 105. doi:10.1186/s12890-020-1141-y

8.	Bediang G, Stoll B, Elia N, Abena J-L, Geissbuhler A. SMS reminders to improve adherence and cure of tuberculosis patients in Cameroon (TB-SMS Cameroon): a randomised controlled trial. BMC Public Health. 2018;18: 583–583.

9.	Hirsch-Moverman Y, Daftary A, Yuengling KA, Saito S, Ntoane M, Frederix K, et al. Using mhealth for HIV/TB treatment support in lesotho: Enhancing patient-provider communication in the start study. J Acquir Immune Defic Syndr. 2017;74: S37–S43.

10.	Das Gupta D, Patel A, Saxena D, Koizumi N, Trivedi P, Patel K, et al. Choice-Based Reminder Cues: Findings From an mHealth Study to Improve Tuberculosis (TB) Treatment Adherence Among the Urban Poor in India. World Med Health Policy. 2020;12: 163–181.

11.	Dessie Gashu K, Nurhussien F, Mamuye A, Alemu Gelaye K, Tilahun B. Developing and Piloting TB Medication and Refilling Reminder System in Ethiopia. Stud Health Technol Inform. 2020;270: 1251–1252. doi:10.3233/SHTI200387

12.	Daftary A, Hirsch-Moverman Y, Kassie G, Melaku Z, Gadisa T, Saito S, et al. A Qualitative Evaluation of the Acceptability of an Interactive Voice Response System to Enhance Adherence to Isoniazid Preventive Therapy Among People Living with HIV in Ethiopia. AIDS Behav. 2017;21: 3057–3067.

13.	Kopanitsa G. A Qualitative Study of the Barriers and Opportunities for Adoption of Web-Portals for Doctors and Patients in Russia. J Med Syst. 2017;41: 62. doi:10.1007/s10916-017-0713-8

14.	Cross A, Gupta N, Liu B, Nair V, Kumar A, Kuttan R, et al. 99DOTS: A Low-Cost Approach to Monitoring and Improving Medication Adherence. Ahmedabad, India: Assoc Computing Machinery; 2019. doi:10.1145/3287098.3287102

15.	Thomas BE, Kumar JV, Onongaya C, Bhatt SN, Galivanche A, Periyasamy M, et al. Explaining Differences in the Acceptability of 99DOTS, a Cell Phone-Based Strategy for Monitoring Adherence to Tuberculosis Medications: Qualitative Study of Patients and Health Care Providers. JMIR MHealth UHealth. 2020;8: e16634.

16.	van den Boogaard J, Lyimo RA, Boeree MJ, Kibiki GS, Aarnoutse RE. Electronic monitoring of treatment adherence and validation of alternative adherence measures in tuberculosis patients: A pilot study. Bull World Health Organ. 2011;89: 632–639.

17.	Ratchakit-Nedsuwan R, Nedsuwan S, Sawadna V, Chaiyasirinroje B, Bupachat S, Ngamwithayapong-Yanai J, et al. Ensuring tuberculosis treatment adherence with a mobile-based CARE-call system in Thailand: a pilot study. Infect Dis Lond Engl. 2020;52: 121‐129. doi:10.1080/23744235.2019.1688862

18.	Bionghi N, Daftary A, Maharaj B, Msibi Z, Amico KR, Friedland G, et al. Pilot evaluation of a second-generation electronic pill box for adherence to Bedaquiline and antiretroviral therapy in drug-resistant TB/HIV co-infected patients in KwaZulu-Natal, South Africa. BMC Infect Dis. 2018;18: 171.

19.	Thomas BE, Kumar JV, Periyasamy M, Khandewale AS, Hephzibah Mercy J, Raj EM, et al. Acceptability of the Medication Event Reminder Monitor for Promoting Adherence to Multidrug-Resistant Tuberculosis Therapy in Two Indian Cities: Qualitative Study of Patients and Health Care Providers. J Med Internet Res. 2021;23: e23294.

20.	Chen SH, Wang I, Hsu HL, Huang CC, Liu YJ, Putri DU, et al. Advantage in privacy protection by using synchronous video observed treatment enhances treatment adherence among patients with latent tuberculosis infection. J Infect Public Health. 2020;13: 1354–1359.

21.	Guo X, Yang Y, Takiff HE, Zhu M, Ma J, Zhong T, et al. A Comprehensive App That Improves Tuberculosis Treatment Management Through Video-Observed Therapy: Usability Study. JMIR MHealth UHealth. 2020;8: e17658.

22.	Guo P, Qiao W, Sun Y, Liu F, Wang C. Telemedicine Technologies and Tuberculosis Management: a Randomized Controlled Trial. Telemed J E Health. 2020;26: 1150‐1156.

23.	Garfein RS, Collins K, Munoz F, Moser K, Cerecer-Callu P, Raab F, et al. Feasibility of tuberculosis treatment monitoring by video directly observed therapy: A binational pilot study. Int J Tuberc Lung Dis. 2015;19: 1057–1064.

24.	Holzman SB, Zenilman A, Shah M. Advancing patient-centered care in tuberculosis management: A mixed-methods appraisal of video directly observed therapy. Open Forum Infect Dis. 2018;5: ofy046. doi:10.1093/ofid/ofy046

25.	Sinkou H, Hurevich H, Rusovich V, Zhylevich L, Falzon D, De Colombani P, et al. Video-observed treatment for tuberculosis patients in Belarus: Findings from the first programmatic experience. Eur Respir J. 2017;49: 1602049.

26.	Navin K, Vadivu G, Maharaj A, Thomas T, Lavanya S, Engn, et al. A Mobile Health Intervention to Support TB Eradication Programme for Adherence to Treatment and a Novel QR Code Based Technique to Monitor Patient-DOTS Provider Interaction. Advanced Computational and Communication Paradigms. Singapore: Springer; 2018. pp. 41–54. Available: ://WOS:000448502300005

27.	Kalita J, Pandey PC, Shukla R, Misra UK. Feasibility and usefulness of tele-follow-up in the patients with tuberculous meningitis. Trans R Soc Trop Med Hyg. 2021;115: 1153–1159. doi:10.1093/trstmh/trab06

28.	Olano-Soler H, Thomas D, Joglar O, Rios K, Torres-Rodriguez M, Duran-Guzman G, et al. Notes from the Field: Use of Asynchronous Video Directly Observed Therapy for Treatment of Tuberculosis and Latent Tuberculosis Infection in a Long-Term-Care Facility - Puerto Rico, 2016-2017. MMWR Morb Mortal Wkly Rep. 2017;66: 1386–1387. doi:10.15585/mmwr.mm6650a5

29.	Iribarren SJ, Sward KA, Beck SL, Pearce PF, Thurston D, Chirico C. Qualitative evaluation of a text messaging intervention to support patients with active tuberculosis: implementation considerations. JMIR MHealth UHealth. 2015;3: e21. doi:10.2196/mhealth.3971

30.	Mahmud N, Rodriguez J, Nesbit J. A text message-based intervention to bridge the healthcare communication gap in the rural developing world. Technol Health Care Off J Eur Soc Eng Med. 2010;18: 137–144. doi:10.3233/THC-2010-0576

31.	Bardosh KL, Murray M, Khaemba AM, Smillie K, Lester R. Operationalizing mHealth to improve patient care: A qualitative implementation science evaluation of the WelTel texting intervention in Canada and Kenya. Glob Health. 2017;13: 87. doi:10.1186/s12992-017-0311-z

32.	Wade VA, Karnon J, Eliott JA, Hiller JE. Home Videophones Improve Direct Observation in Tuberculosis Treatment: A Mixed Methods Evaluation. PLoS ONE. 2012;7: e50155. doi:10.1371/journal.pone.0050155

33.	Gashu KD, Gelaye KA, Tilahun B. Feasibility, acceptability and challenges of phone reminder system implementation for tuberculosis pill refilling and medication in Northwest Ethiopia. Res Sq. 2021; Version 1. doi:10.21203/rs.3.rs-229284/v1

34.	Mohammed S, Siddiqi O, Ali O, Habib A, Haqqi F, Kausar M, et al. User engagement with and attitudes towards an interactive SMS reminder system for patients with tuberculosis. J Telemed Telecare. 2012;18: 404–408. doi:10.1258/jtt.2012.120311

35.	Hermans SM, Elbireer S, Tibakabikoba H, Hoefman BJ, Manabe YC. Text messaging to decrease tuberculosis treatment attrition in TB-HIV coinfection in Uganda. Patient Prefer Adherence. 2017;11: 1479–1487. doi:10.2147/PPA.S135540

36.	Browne SH, Umlauf A, Tucker AJ, Low J, Moser K, Garcia JG, et al. Wirelessly observed therapy compared to directly observed therapy to confirm and support tuberculosis treatment adherence: A randomized controlled trial. PLoS Med. 2019;16: e1002891.

37.	Bendiksen R, Ovesen T, Asfeldt AM, Halvorsen DS, Gravningen K. Use of video call in the treatment of tuberculosis disease in Northern Norway. Tidsskr Nor Laegeforen. 2020;140: 35–40. doi:10.4045/tidsskr.19.0322

38.	DeMaio J, Schwartz L, Cooley P, Tice A. The application of telemedicine technology to a directly observed therapy program for tuberculosis: A pilot project. Clin Infect Dis. 2001;33: 2082–2084. doi:10.1086/324506

39.	Gassanov MA, Feldman LJ, Sebastian A, Kraguljac MJ, Rea E, Yaffe B. The use of videophone for directly observed therapy for the treatment of tuberculosis. Can J Public Health. 2013;104: e272.

40.	Story A, Aldridge RW, Smith CM, Garber E, Hall J, Ferenando G, et al. Smartphone-enabled video-observed versus directly observed treatment for tuberculosis: a multicentre, analyst-blinded, randomised, controlled superiority trial. The Lancet. 2019;393: 1216–1224. doi:10.1016/S0140-6736(18)32993-3

41.	Ting NCH, El-Turk N, Chou MSH, Dobler CC. Patient-perceived treatment burden of tuberculosis treatment. PLoS ONE. 2020;15: e0241124.

42.	Wade V, Izzo J, Hamlyn J. Videophone delivery of Medication Management in Community Nursing. Electron J Health Inform. 2009;4: 5. doi:10.1007/8754_2011_26

43.	Belknap R, Weis S, Brookens A, Au-Yeung KY, Moon G, DiCarlo L, et al. Feasibility of an Ingestible Sensor-Based System for Monitoring Adherence to Tuberculosis Therapy. PLoS ONE. 2013;8: e53373.

44.	Garfein RS, Liu L, Cuevas-Mota J, Collins K, Catanzaro DG, Munoz F, et al. Evaluation of recorded video-observed therapy for anti-tuberculosis treatment. Int J Tuberc Lung Dis. 2020;24: 520–525.

45.	Bommakanti KK, Smith LL, Liu L, Do D, Cuevas-Mota J, Collins K, et al. Requiring smartphone ownership for mHealth interventions: who could be left out? BMC Public Health. 2020;20: 81.

46.	Burzynski J, Mangan JM, Lam CK, Macaraig M, Salerno MM, deCastro BR, et al. In-Person vs Electronic Directly Observed Therapy for Tuberculosis Treatment Adherence: A Randomized Noninferiority Trial. JAMA Netw Open. 2022;5: e2144210.

47.	Horter S, Stringer B, Venis S, Du Cros P. “I can also serve as an inspiration”: A qualitative study of the TB&Me blogging experience and its role in MDRTB treatment. PLoS ONE. 2014;9: e108591.

48.	Bassett IV, Coleman SM, Giddy J, Bogart LM, Chaisson CE, Ross D, et al. Sizanani: a Randomized Trial of Health System Navigators to Improve Linkage to HIV and TB Care in South Africa. J Acquir Immune Defic Syndr. 2016;73: 154‐160. doi:10.1097/QAI.0000000000001025

49.	Liu X, Lewis JJ, Zhang H, Lu W, Zhang S, Zheng G, et al. Effectiveness of Electronic Reminders to Improve Medication Adherence in Tuberculosis Patients: A Cluster-Randomised Trial. PLoS Med. 2015;12: e1001876. doi:10.1371/journal.pmed.1001876

50.	Thekkur P, Kumar AMV, Chinnakali P, Selvaraju S, Bairy R, Singh AR, et al. Outcomes and implementation challenges of using daily treatment regimens with an innovative adherence support tool among HIV-infected tuberculosis patients in Karnataka, India: a mixed-methods study. Glob Health Action. 2019;12: 1–11. doi:10.1080/16549716.2019.1568826

51.	Prabhu A, Agarwal U, Singla N, Sarin R, Tripathy JP, Sagili K, et al. “99DOTS” techno-supervision for tuberculosis treatment - a boon or a bane? Exploring challenges in its implementation at a tertiary centre in Delhi, India. Indian J Tuberc. 2020;67: 46–53. doi:10.1016/j.ijtb.2019.08.010

52.	Drabarek D, Anh NT, Nhung NV, Hoa NB, Fox GJ, Bernays S. Implementation of Medication Event Reminder Monitors among patients diagnosed with drug susceptible tuberculosis in rural Viet Nam: a qualitative study. PLoS ONE. 2019;14: e0219891. doi:10.1371/journal.pone.0219891

53.	Buchman T, Cabello C. A New Method to Directly Observe Tuberculosis Treatment: Skype Observed Therapy, a Patient-Centered Approach. J Public Health Manag Pract. 2017;23: 175–177.

54.	Cox SN, Elf J, Lokhande R, Ogale YP, DiAndreth L, Dupuis E, et al. Mobile phone access and comfort: Implications for HIV and tuberculosis care in India and South Africa. Open Forum Infect Dis. 2018;5: S162.

55.	Chuck C, Robinson E, Macaraig M, Alexander M, Burzynski J. Enhancing management of tuberculosis treatment with video directly observed therapy in New York City. Int J Tuberc Lung Dis. 2016;20: 588–593.

56.	Holzman SB, Atre S, Sahasrabudhe T, Ambike S, Jagtap D, Sayyad Y, et al. Use of Smartphone-Based Video Directly Observed Therapy (vDOT) in Tuberculosis Care: Single-Arm, Prospective Feasibility Study. JMIR Form Res. 2019;3: e13411. doi:10.2196/13411

57.	Stagg HR, Lewis JJ, Liu X, Huan S, Jiang S, Chin DP, et al. Temporal Factors and Missed Doses of Tuberculosis Treatment. A Causal Associations Approach to Analyses of Digital Adherence Data. Ann Am Thorac Soc. 2020;17: 438‐449. doi:10.1513/AnnalsATS.201905-394OC

58.	Trajman A, Long R, Zylberberg D, Dion MJ, Al-Otaibi B, Menzies D. Factors associated with treatment adherence in a randomised trial of latent tuberculosis infection treatment. Int J Tuberc Lung Dis. 2010;14: 551‐559.

59.	Lam CK, McGinnis Pilote K, Haque A, Burzynski J, Chuck C, Macaraig M. Using Video Technology to Increase Treatment Completion for Patients With Latent Tuberculosis Infection on 3-Month Isoniazid and Rifapentine: An Implementation Study. J Med Internet Res. 2018;20: e287. doi:10.2196/jmir.9825

60.	Mohammed S, Glennerster R, Khan AJ. Impact of a Daily SMS Medication Reminder System on Tuberculosis Treatment Outcomes: a Randomized Controlled Trial. PLoS ONE. 2016;11: e0162944. doi:10.1371/journal.pone.0162944

61.	Person AK, Blain ML, Jiang H, Rasmussen PW, Stout JE. Text messaging for enhancement of testing and treatment for tuberculosis, human immunodeficiency virus, and syphilis: a survey of attitudes toward cellular phones and healthcare. Telemed J E Health. 2011;17: 189–195. doi:10.1089/tmj.2010.0164
